# Randomized trials of ‘personalized’, ‘individualized’ and ‘precision’ interventions are very diverse and have low transparency and high bias

**DOI:** 10.64898/2026.02.09.26345904

**Authors:** Luigi Russo, Nicolò Lentini, Luisa Soru, Roberta Pastorino, Stefania Boccia, John PA Ioannidis

**Author notes:** **Corresponding Author**: Nicolò Lentini MSc, Mail, Section of Hygiene, University Department of Life Sciences and Public Health, Università Cattolica del Sacro Cuore, 00168 Rome, Italy.

## Abstract

**Background and objective:** The terms ‘personalized’, individualized’ and ‘precision’ are increasingly used to describe interventions, yet their operational meaning is often unclear. Despite extensive conceptual debate, no empirical work has examined how these terms are applied in randomized controlled trials (RCTs) and how transparent and reliable the results of these trials are.

**Methods:** We surveyed MEDLINE for RCTs published between 2020 and 2022 that used the terms ‘personalized’, ‘individualized’, or ‘precision’ in their title to describe an intervention. We collected data on trial characteristics and conclusions, intervention features, individual-level tailoring, transparency indicators, and risk of bias. Interventions were classified according to the number of personalized components, modes of tailoring, and dosage variability.

**Results:** A total of 262 RCTs were included. The term ‘personalized’ was used most frequently (n=129, 49.2%), followed by ‘individualized’ (n=120, 45.8%) and ‘precision’ (n=13, 5.0%). Most trials compared personalized interventions versus non-personalized control groups (n=225, 85.9%) and focused on therapeutic applications (n=186, 71.0%). Behavioral, digital, and medication-based interventions predominated. Personalization was most often based on lifestyle, psychological characteristics, or disease classification, whereas genetic and omics-based features were relatively uncommon. The majority of trials implemented a single intervention (n=177, 67.6%) tailored differently for each participant (n=217, 82.8%), often through individualized dosage or content (n=109, 41.6%). Of the 221 RCTs comparing personalized interventions versus non-personalized controls, abstract conclusions were favorable to the personalized intervention in 156 (70.6%), mixed in 36 (16.3%) and unfavorable in 29 (13.1%). Few meaningful differences were observed across studies according to the term used. Transparency indicators such as data (n=13, 5.0%) and code sharing (n=1, 0.4%) were rare, and most trials (n=162, 68.6%) were judged to be at high overall risk of bias.

**Conclusions:** In contemporary RCTs, the labels ‘personalized’, ‘individualized’, and ‘precision’ are applied largely interchangeably to a wide range of heterogeneous interventions that are predominantly non-genomic. Most of these trials have favorable conclusions, low transparency and high risk of bias.

**What is new?:**

1. Key findings
  - In a large sample of 262 randomized trials, the terms “personalized,” “individualized,” and “precision” were used largely interchangeably despite substantial conceptual differences.
  - Most interventions labeled as personalized relied on behavioral, clinical, or contextual features rather than genetic or omics information.
2. What this adds to what is known?
  - The vast majority of trials reported favorable conclusions while frequently showing high risk of bias and limited transparency practices.
  - This study provides the first large-scale empirical assessment linking the use of personalization labels with methodological rigor and transparency.
3. **What is the implication, what should change now?**
  - Greater conceptual clarity, improved reporting of tailoring methods, and stronger transparency standards are needed to ensure the credibility of personalized intervention research.

## Introduction

Personalized, individualized, and precision medicine represent a rapidly evolving and frequently touted domains within healthcare. These terms are commonly used to describe an approach that tailors prevention, diagnosis, and treatment to individual characteristics, including genetic, environmental, and lifestyle factors. Multiple definitions have been proposed from authoritative bodies. For example, the National Human Genome Research Institute [1] defines personalized medicine narrowly, as the use of an individual’s genetic profile to guide disease prevention, diagnosis, and treatment. In contrast, the International Consortium for Personalized Medicine (ICPerMed) [2] adopts a broader perspective, viewing it as a model that integrates both phenotypic and genotypic information (e.g., molecular profiling, imaging, lifestyle data) to inform therapy or prevention. Despite this shared overarching principle, the interpretation of the terms ‘personalized’, ‘individualized’, and ‘precision’ varies widely across disciplines and is often inconsistent even within the same field, as illustrated by the differing definitions above.

These concepts have permeated numerous medical specialties, illustrating their versatility but also the lack of consensus on their exact meaning. In oncology, for example, precision medicine often refers to genetic profiling that identifies mutations and guides targeted therapies [3,4]. In cardiology, personalized risk assessments combining genetic and lifestyle factors aim to inform individualized prevention strategies, such as interventions based on lifetime genetic risk scores [5,6]. Beyond genetics, the term ‘personalized’ has been applied to very diverse interventions. Illustratively, these include tailored physiotherapy after surgery, customized text-message reminders for adherence [7], and music therapy for dementia patients [8].

The existing literature reflects this diversity of interpretation. Editorials, commentaries, and reviews have examined, debated, and promoted (often with some hype) the origins, evolution, and future directions of personalized, individualized and precision medicine. Broadly, these writings fall into two groups: those that conceive of the concepts as an integration of genetic, social, and environmental features [9], and those that focus primarily on biological and genomic applications [10]. Ethical and social concerns [11], particularly related to privacy and data use, equity in access, and the use of genetic or other sensitive individual-level information. along with differences in public understanding [12], add further complexity. Despite the growing use of these terms in published clinical trials, the way they are used in practice remains unclear. Different articles may emphasize definitional debates [13], argue over which features should be included, or speculate about future developments [14,15], while the real-world implications of the terminology remain underexplored.

Clarifying how these terms are applied in clinical research is therefore essential. To address this gap, we surveyed randomized controlled trials (RCTs) published between 2020 and 2022 that describe interventions as ‘personalized’, ‘individualized’, or ‘precision’. The aim was to empirically examine how these terms are used in randomized trials by characterizing the features of interventions labeled as such and analyzing how personalization is operationalized across different clinical contexts. Having assembled this literature corpus, we also aimed to assess whether these trials are transparent and whether they have low or high risk of bias.

## Methods

We previously registered the protocol for this study on Open Science Framework [16]. The protocol aimed to generate a literature corpus of RCTs that prominently use in their titles the terms of interest to describe assessed interventions. Once the eligible trials were assembled and we could have a better sense of what they pertained to, we further extended the protocol to characterize better these trials and to explore features of transparency and bias. The full list of trials in available in Supplementary File 1 for further methodological research.

### Search strategy

We searched for articles published between 2020 and 2022, in English, and indexed in Medline. We focused on RCTs that used the terms of interest to prominently describe the intervention in their title. We applied the following search string:

(precision [ti] OR personalized [ti] OR personalised [ti] OR individualized [ti] OR individualised [ti]) Filter[RandomizedControlledTrial]

After conducting the literature search, we evaluated each article against the following eligibility criteria: (1) any RCT on humans, including parallel arm, cross-over, cluster randomized or any other randomized design; and (2) randomization of the participants to an intervention considered ‘personalized’, ‘individualized’, or ‘precision’. Both articles reporting the results of RCTs and those reporting protocols of RCTs were deemed eligible.

We excluded articles that did not report either the protocol or the results of an RCT (e.g., reviews, editorials, perspectives). We also excluded articles in which the relevant terms were not used to describe a randomized intervention, but rather for other purposes, as well as articles that did not address an actual or possible clinical condition and those not related to medicine and health.

### Screening

Identified articles were uploaded into the Rayyan software. Two reviewers (L.R., A.P.) independently performed screening based on title and abstract, and disagreements were resolved through consensus. Full texts of the identified articles were retrieved and reviewed independently by two researchers (L.R., L.S.), and disagreements were resolved by consensus. Articles meeting the eligibility criteria were subsequently entered into data extraction.

### Data extraction

Two reviewers (L.R., L.S.) independently performed data extraction. From each included randomized controlled trial, we extracted:

*General information*: title, first author, DOI, journal of publication, year of publication, country, type of paper (protocol, trial).

*Study characteristics*: medical specialty (according to European Union directive [17]), specific disease, number of arms, total sample size, and study group. Studies were classified into two main categories:

Group 1: the personalized/individualized/precision intervention is compared to a non-personalized/individualized/precision intervention. This group was further subdivided post hoc into:

⍰ Group 1a: studies with two or more intervention arms but only one personalized/individualized/precision;

⍰ Group 1b: studies including more than one personalized/individualized/precision intervention arms

Group 2: the personalized/individualized/precision intervention is compared to another personalized/individualized/precision intervention.

Furthermore, for articles presenting trial results, we extracted whether the abstract conclusion was favorable to the personalized intervention, had mixed conclusions, or was unfavorable compared with the control. For articles in group 2 and for protocols this classification was not applicable.

*Personalized, individualized, or precision intervention characteristics*: we extracted which term was used; the type of application (i.e., primary prevention, secondary prevention, therapy); and the type of intervention (digital, omics, behavioral, dietary, medication, surgery, physical exercises, radiotherapy, anesthesia, physiotherapy, rehabilitation, psychological). We evaluated whether the intervention provided was the same for all participants in the intervention arm; if it was not, we extracted how the intervention differed across participants in its delivery (schedule of therapy, dosage of therapy, follow-up/visits, testing, drug used, anatomical target (e.g., radiotherapy applied to different body areas/organs; prostheses applied to different parts of the same bone or tissue). For dosage or schedule of therapy, we considered not only pharmacological treatments but any type of intervention, including, for example, diets with different caloric intake or different intensity in physical activity.

We extracted a description of the intervention, including the specific features used to personalize it, the category of individual features on which the intervention was based (as reported below), and the control arm intervention.

The individual features upon which the intervention is personalized were classified in the following main categories and subcategories: demographics (age, sex, place of birth, domicile), socioeconomic (financial status, cultural and social, education), anthropometrics (height, weight, BMI), genetic (genetic testing, monogenic disorder), lifetime risk, omics (microbiome, polygenic risk score, metabolomics, nutrigenomics, proteomics), lifestyle and behavioral, psychological (patient preferences, motivation), vital signs (blood pressure, temperature, pulse rate, respiration rate), imaging tools (radiologic, magnetic resonance, scintigraphy, ECG, PET, ultrasound, etc.), tests on biological material (blood, urine, biopsy, other), complex scores (specify), physical or functional capacity, disease classification or other (specify).

As part of post hoc data extraction, we extracted from each study the number of personalized/individualized/precision interventions present and classified them as 1, 2, 3, 4, or multiple (>4). Moreover, we extracted from each study how the intervention content was tailored across participants, defined as the number of different ways in which the personalized intervention was delivered, and classified this as 1, 2, 3, 4, multiple (>4), or different for each patient. Finally, for studies that included differences in dosage, we extracted the number of dosage levels present (1, 2, 3, 4, multiple [>4], or for each patient), whereas studies without dosage differences were recorded as “no dose involved.” This approach allowed us to capture studies in which a single personalized intervention (e.g., a diet) was conceptually one intervention but varied at the individual level in terms of content, dose, or both.

Data extraction was performed by two independent researchers (L.R., L.S.), with discrepancies discussed for consensus or settled by a third assessor (N.L.).

### Transparency indicators

For articles describing trial results, transparency indicators were assessed through both manual data extraction and the application of the previously validated, publicly available *Rtransparent* algorithm [18] which identifies code sharing, data sharing, trial registration, disclosures of conflicts of interest, and funding disclosures. The algorithm has been shown to have high specificity (>98%) and generally high sensitivity (>76%) [18]. Any discrepancies between manual extraction and algorithmic results were resolved by a third reviewer (N.L.). For registered trials, we extracted whether the registration was prospective, retrospective or before completion.

### Risk of Bias assessment

To assess the risk of bias of the included studies we utilized the revised Cochrane Risk-of-Bias Tool for Randomized Trials (RoB 2) [19], through the ROBoto2 webapp [20], an open-source, web-based platform for large language model (LLM)-assisted risk of bias assessment of clinical trials. All LLM-generated outputs were reviewed and verified by human experts (L.R.), who could accept or modify the model’s answers, ensuring accuracy. ROBoto2 has been validated in previous studies, showing high specificity and moderate sensitivity [20]. For graphical visualization, we used the Risk-of-bias VISualization (robvis) webapp [21].

### Data synthesis

The results were described narratively and through tables and graphs, where appropriate. Descriptive statistics were presented for the characteristics of the eligible studies. Trial characteristics and main outcomes were reported as counts, percentages for categorical variables, and medians with interquartile ranges (IQRs) for continuous variables.

As post hoc synthesis, we classified each study based on the combination of number of personalized/individualized/precision intervention(s) and the way of tailoring it, and the number of different doses. For example, a study with personalized feedback was classified as a single intervention, with no dose involved, and tailored differently for each participant.

### Statistical analysis

Exploratory analyses were conducted to assess whether there were differences in the characteristics and features of articles depending on whether they were (1) protocols or results; (2) used the term ‘personaliz(s)ed’, ‘individualiz(s)ed’, or ‘precision’; and (3) had favorable conclusions for the personalized intervention (among those that had results rather than just protocols). We used Pearson’s χ² or Fisher’s exact tests to assess whether each category differed between type of study, term used and conclusions of the abstract. For associations between ordinal categorical variables with more than two levels, we applied a linear-by-linear association test (Mantel test for trend) to account for the ordered structure of the data. For continuous variables, the Kruskal–Wallis test or the Mann–Whitney U test was applied, as appropriate.

Agreement between *Rtransparent* automated algorithm extraction and human extraction for transparency-related fields was assessed using Cohen’s kappa coefficient. For discordant cases, discrepancies were reviewed to determine the correct classification.

A 2-sided P < .005 was considered statistically significant, as a conservative threshold to account for multiple comparisons given that the exact number of tests was not pre-specified. P < .05 was considered suggestive [22]. All analyses were performed using R version 4.4.0 (2024-04-24) for Windows.

## Results

A total of 435 records were identified and screened by title and abstract. Of these, 313 articles were assessed for full-text eligibility, and 262 were finally included in the review. Fifty-five articles were excluded after full-text assessment for the following reasons: ineligible intervention either non-medical condition (n=8) or non-personalized intervention (n = 3); article not possible to retrieve (n = 10); unsuitable study design including secondary analysis (n=11), non-randomized design (n=10), sub-study of an RCT (n=4), economic evaluation (n=1); non-English language (n = 2); and retracted article (n = 2). Figure 1 shows the PRISMA flowchart.

**Figure 1.**
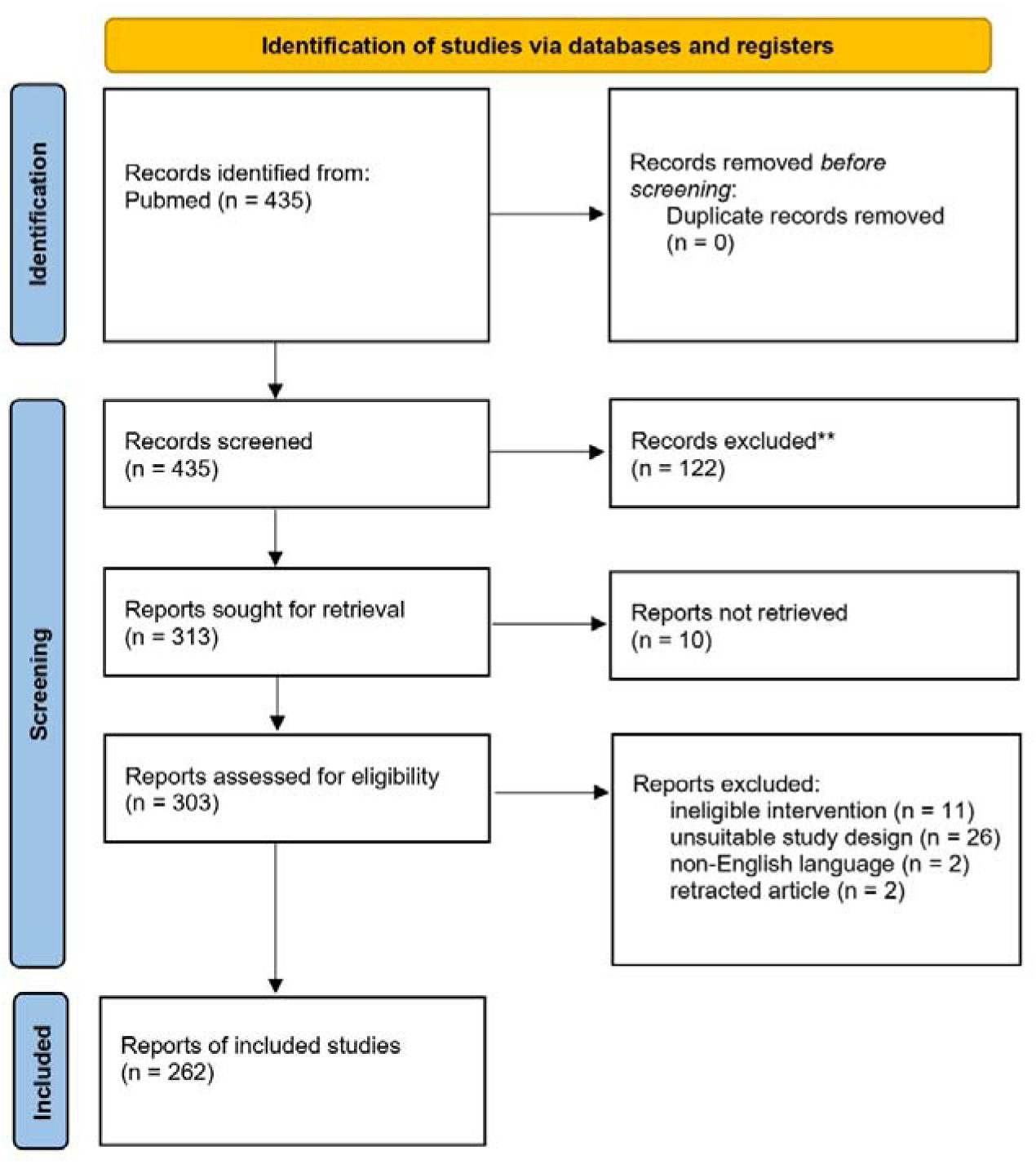
PRISMA flow diagram of the study search and selection process.

### Characteristics of the included trials

Among the 262 included trials (Table 1), the term ‘personalized’ was used most frequently (n=129; 49.2%), followed by ‘individualized’ (n=120; 45.8%) while ‘precision’ was uncommon (n=13; 5.0%). The majority were reports of trial results but the corpus included also 28 protocols. Geographic location was highly diverse. Most trials had two arms (n=221; 84.4%), while 32 (12.2%) and 8 (3.1%) had three and four arms, respectively, while one had 6 arms. The median sample size was 136 (IQR 63.3–302.3). Studies were largely classified in group 1a (n=225; 85.9%) and group 1b (n=21; 8.0%). Among 218 RCTs that compared personalized interventions versus non-personalized controls, the conclusion of the Abstract was favorable to the personalized/individualized/precision intervention in 156 (70.6%), mixed in 36 (16.3%), and unfavorable in only 29 (13.1%).

**Table 1.**
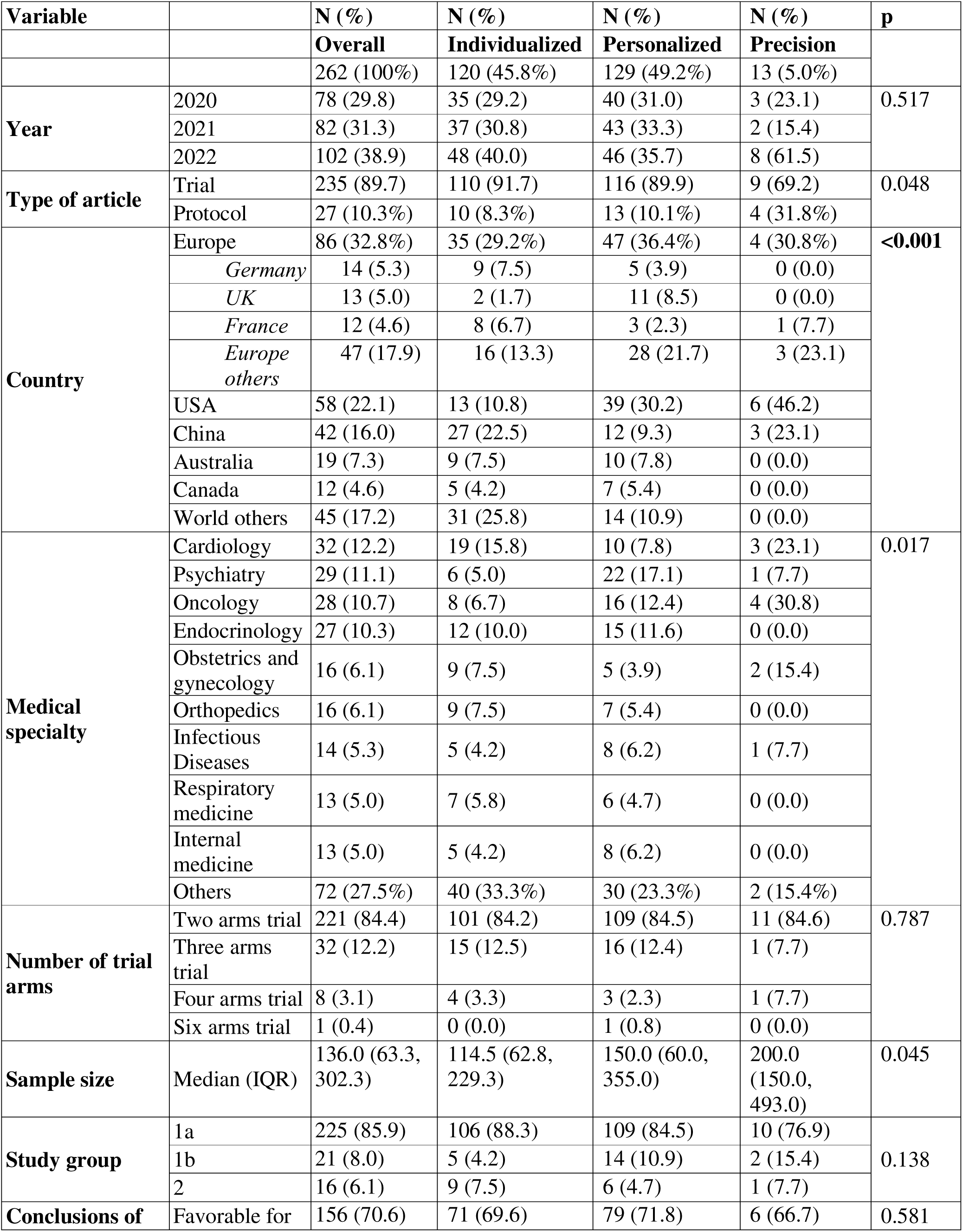
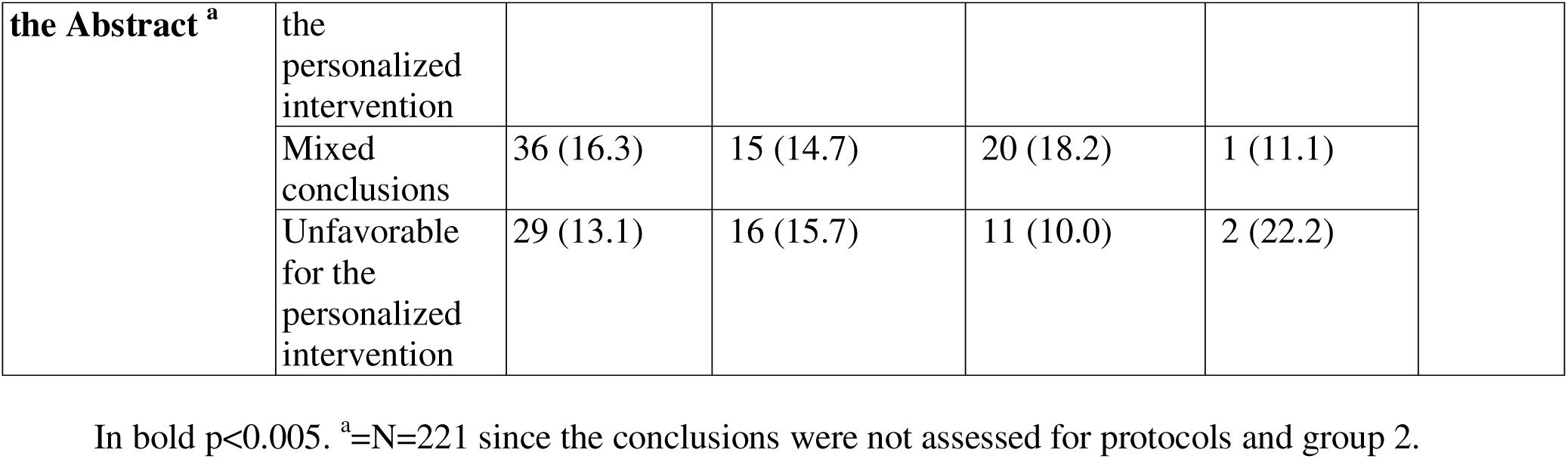
Characteristics of the eligible RCTs.

### Characteristics of the intervention

As shown in Table 2, more than two-thirds of the interventions were therapeutic rather than preventive. Intervention types varied widely, with the most common being behavioral approaches (n=87; 33.2%), digital interventions (n=77; 29.4%) and medication-based interventions (n=72; 27.5%). Within the experimental arm, differences most frequently pertained to the dosage of therapy (n=136; 51.9%). A total of 573 individual features were identified as the main categories on which personalization was based. Within this vast variety of features, lifestyle and behavioral features (n=98; 37.4%), classification of the disease (n=75; 28.6%), and psychological features (n=71; 27.1%), were the most common, while genetic and omics features were uncommon. Most studies employed a single intervention (n=177; 67.6%), and most studies tailored the intervention(s) to each patient (n=217; 82.8%). In those with dosage variations, the dosage most often differed for each patient (n=109; 41.6%). Hence, the most common combinations were one intervention with both dosage and tailoring different for each patient (n=44; 36.7%), and one intervention, with tailoring different for each patient and no dose involved (n=29; 24.2%) (Table 3).

**Table 2.**
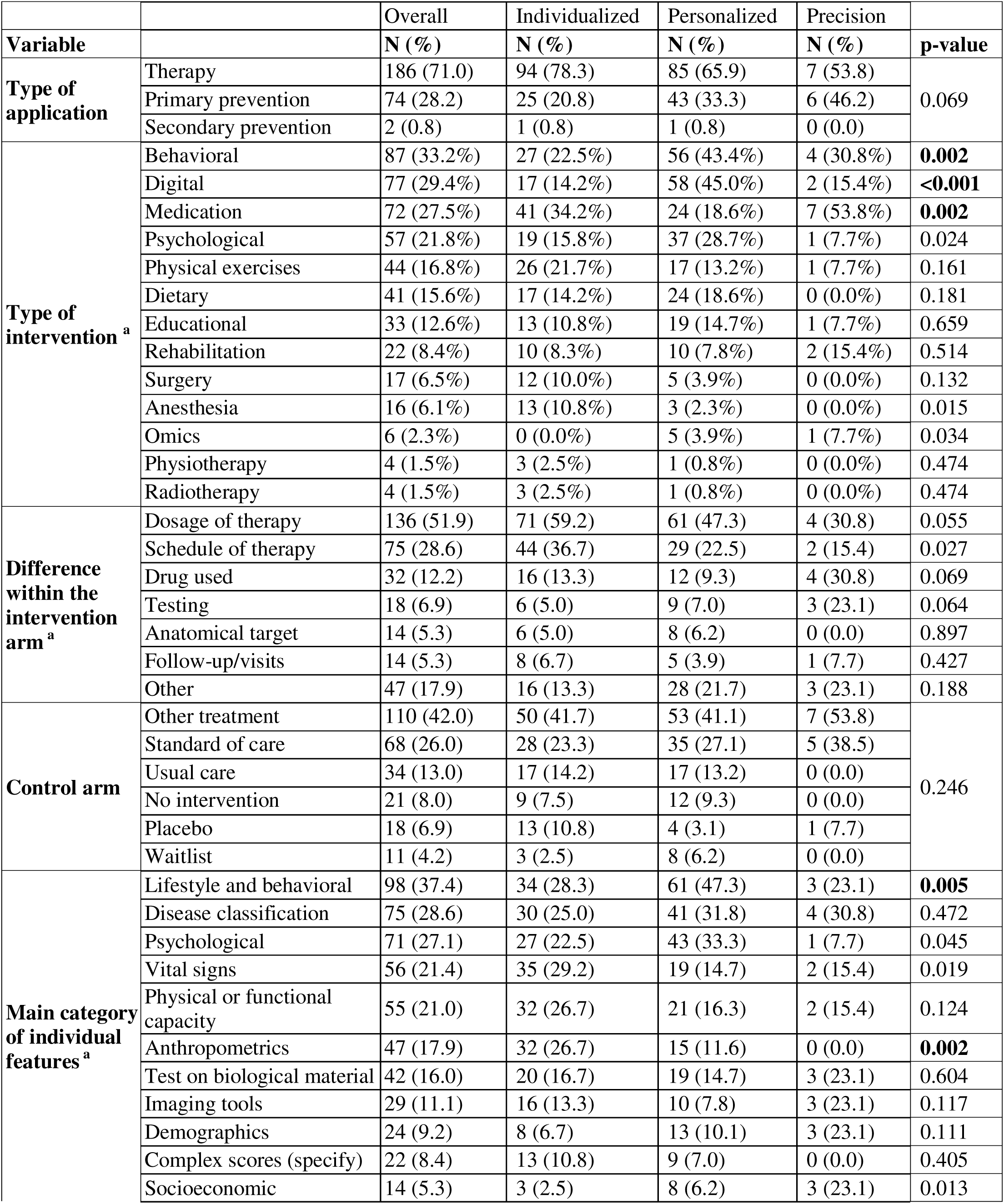
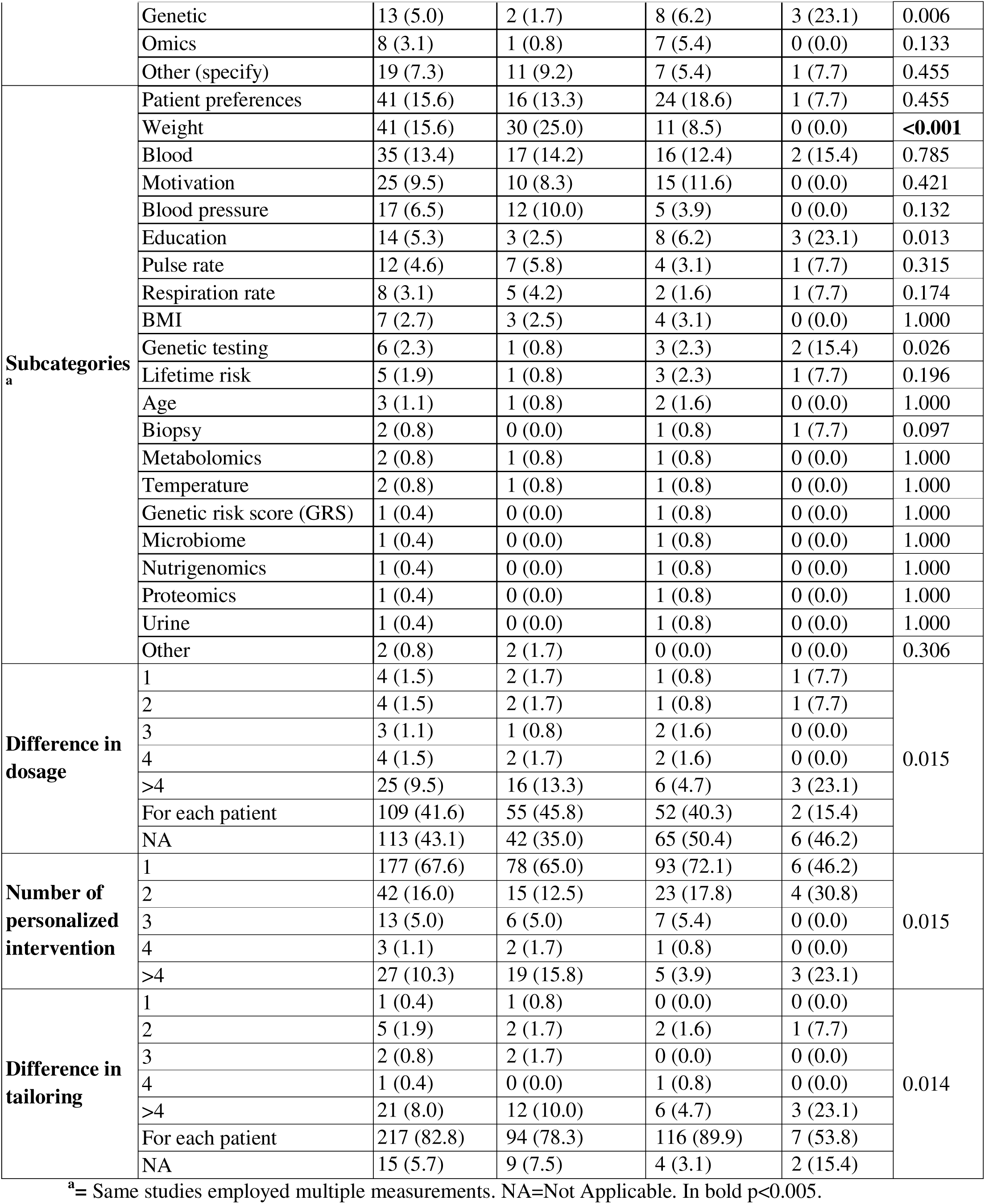
Characteristics of the personalized, individualized, or precision intervention in eligible trials.

**Table 3.** Combination of number of intervention(s), number of dosage(s) and tailoring of the intervention(s).

| Combination in the intervention |  |  | Term used |  |  | Conclusion of the abstract |  |  | Type of study |  |
| --- | --- | --- | --- | --- | --- | --- | --- | --- | --- | --- |
| Number of interventions | Tailoring | Dosage | Individualized (N (%)) | Personalized (N (%)) | Precision (N (%)) | Favorable (N (%)) | Mixed (N (%)) | Unfavorable (N (%)) | Protocol (N (%)) | Trial (N (%)) |
| One | One | No dose | 1 (0.8) | - | - | 1 (0.6) | - | - | - | 1 (0.4) |
|  | Two | Multiple (>4) | 1 (0.8) | - | - | 1 (0.6) | - | - | - | 1 (0.4) |
|  | Four | Four | - | 1 (0.8) | - | 1 (0.6) | - | - | - | 1 (0.4) |
|  | Multiple (>4) | Multiple (>4) | - | 1 (0.8) | 1 (7.7) | - | 1 (2.8) | - | 1 (3.7) | 1 (0.4) |
|  | Different for each | No dose | 29 (24.2) | 47 (36.4) | 2 (15.4) | 46 (29.5) | 14 (38.9) | 10 (34.5) | 8 (29.6) | 70 (29.8) |
|  |  | Different for each | 44 (36.7) | 43 (33.3) | 2 (15.4) | 58 (37.2) | 9 (25.0) | 11 (37.9) | 9 (33.3) | 80 (34.0) |
|  | NA | Fixed | 2 (1.7) | 1 (0.8) | 1 (7.7) | 3 (1.9) | - | 1 (3.4) | - | 4 (1.7) |
|  |  | Two | 1 (0.8) | - | - | 1 (0.6) | - | - | - | 1 (0.4) |
|  |  | Multiple (>4) | 1 (0.8) | - | - | - | - | 1 (3.4) | - | 1 (0.4) |
| Two | Two | No dose | - | 2 (1.6) | 1 (7.7) | 3 (1.9) | - | - | - | 3 (1.3) |
|  |  | Multiple (>4) | 1 (0.8) | - | - | - | - | - | - | 1 (0.4) |
|  | Multiple (>4) | Multiple (>4) | 1 (0.8) | 3 (2.3) | 1 (7.7) | 2 (1.3) | 1 (2.8) | - | 1 (3.7) | 4 (1.7) |
|  | Different for each | No dose | 5 (4.2) | 10 (7.8) | 1 (7.7) | 7 (4.5) | 2 (5.6) | 1 (3.4) | 1 (3.7) | 15 (6.4) |
|  |  | Different for each | 6 (5.0) | 7 (5.4) | - | 6 (3.8) | 1 (2.8) | - | 3 (11.1) | 10 (4.3) |
|  | NA | Fixed | 1 (0.8) | 1 (0.8) | 1 (7.7) | 2 (1.3) | - | - | 1 (3.7) | 2 (0.9) |
| Three | Three | No dose | 2 (1.7) | - | - | 2 (1.3) | - | - | - | 2 (0.9) |
|  | Multiple (>4) | Multiple (>4) | 1 (0.8) | - | - | - | - | - | - | 1 (0.4) |
|  | Different for each | No dose | - | 3 (2.3) | - | - | 2 (5.6) | - | - | 3 (1.3) |
|  |  | Different for each | 3 (2.5) | 1 (0.8) | - | 4 (2.6) | - | - | - | 4 (1.7) |
|  | NA | Fixed | 1 (0.8) | 2 (1.6) | - | 3 (1.9) | - | - | - | 3 (1.3) |
| Four | Different for each | Different for each | 1 (0.8) | - | - | - | 1 (2.8) | - | - | 1 (0.4) |
|  | NA | Fixed | 1 (0.8) | - | - | 1 (0.6) | - | - | - | 1 (0.4) |
| Multiple (>4) | Multiple (>4) | Multiple (>4) | 12 (10.0) | 2 (1.6) | 1 (7.7) | 7 (4.5) | 3 (8.3) | 5 (17.2) | - | 15 (6.4) |
|  | Different for each | No dose | 5 (4.1) | 3 (2.3) | 2 (15.4) | 8 (5.1) | 1 (2.8) | - | 1 (3.7) | 9 (3.8) |
|  |  | Multiple (>4) | - | 1 (0.8) | - | - | - | - | 1 (3.7) | - |
|  |  | Different for each | 1 (0.8) | 1 (0.8) | - | - | 1 (2.8) | - | 1 (3.7) | 1 (0.4) |

### Association analyses

The association analysis revealed almost no differences in applications and features among studies employing different terms (Table 2), having different abstract conclusions (Supplementary Table 1) or studies published versus protocols (Supplementary Table 2). The few significant patterns observed were that digital and behavioral interventions almost always were termed ‘personalized’ while medication interventions were termed ‘individualized’, studies with interventions tailored on lifestyle and behavioral employed more commonly ‘personalized’ while those tailored on anthropometrics features were more frequently ‘individualized’. Finally, studies on weight outcomes were largely termed ‘individualized’ (Supplementary Table 3).

### Transparency among trials

Statements on code sharing were rare, with only 1 study (0.4%) providing open code. Similarly, open data availability was rare (n=13, 5.1%). On the other hand, presence of statements on funding source (n=244; 93.4%) and on conflict of interest (n=244; 93.4%) were present in almost all of the studies. 202 studies (78.3%) were registered in a trial repository, but of those only 155 (59.1%) were registered prospectively.

Comparison between the *Rtransparent* automated extraction and human assessment revealed moderate agreement for funding declarations (κ = 0.59) and conflict of interest declarations (κ = 0.51), and lower agreement for study registration (κ = 0.39). Agreement was more substantial for data availability (κ = 0.63). Open code availability was proposed in 2 cases by *Rtransparent*, and 2 cases by manual extraction (with none overlapping) (κ = −0.01) (Table 4).

**Table 4.** Comparison of transparency-related patterns across studies assessed by a human reviewer and the *Rtransparent* algorithm.

| Variable | N (%) | Kappa | Conflicts in assessment | Accurate extraction |  |
| --- | --- | --- | --- | --- | --- |
|  |  |  |  | Human | *Rtransparent* |
| Declaration of Funding | 244 (93.1) | 0.541 | 21 (8.0%) | 9 (42.9%) | 12 (57.1%) |
| Declaration of Conflict of interest | 245 (93.5) | 0.508 | 11 (4.2%) | 2 (18.2%) | 9 (81.8%) |
| Registration on a trial repository | 204 (77.9) | 0.370 | 44 (16.8%) | 41 (93.2%) | 3 (6.8%) |
| *Before completion* | 25 (12.3) |  |  |  |  |
| *Not known* | 2 (1.0) |  |  |  |  |
| *Retrospectively* | 22 (10.8) |  |  |  |  |
| *Prospectively* | 155 (76.0) |  |  |  |  |
| Data availability |  | 0.640 | 46 (17.6%) | 29 (63.0%) | 17 (37.0%) |
| *Data fully available* | 13 (5.0) | - | - | - | - |
| *Data not available* | 171 (65.3) | - | - | - | - |
| *Data available upon request* | 78 (29.8) | - | - | - | - |
| Code availability |  | -0.008 | 4 (1.5%) | 3 (75.0%) | 1 (25.0%) |
| *Code available* | 1 (0.4) | - | - | - | - |
| *Code not available* | 257 (98.1) | - | - | - | - |
| *Code available upon request* | 4 (1.5) | - | - | - | - |

### Risk of bias assessment

The results of the risk of bias evaluation are reported in Figure 2 and in Supplementary File 2. Overall, 162 studies (68.6%) were assessed as being at high risk of overall bias, 32 (13.6%) raised some concerns, and 42 (17.8%) were judged to be at low risk. The domains most frequently contributing to a high risk of bias were bias in selection of the reported result (n=126; 53.4%), bias in measurement of the outcome (n=83; 35.2%), and bias arising from the randomization process (n=79; 33.5%) (Supplementary Table 4).

**Figure 2.**
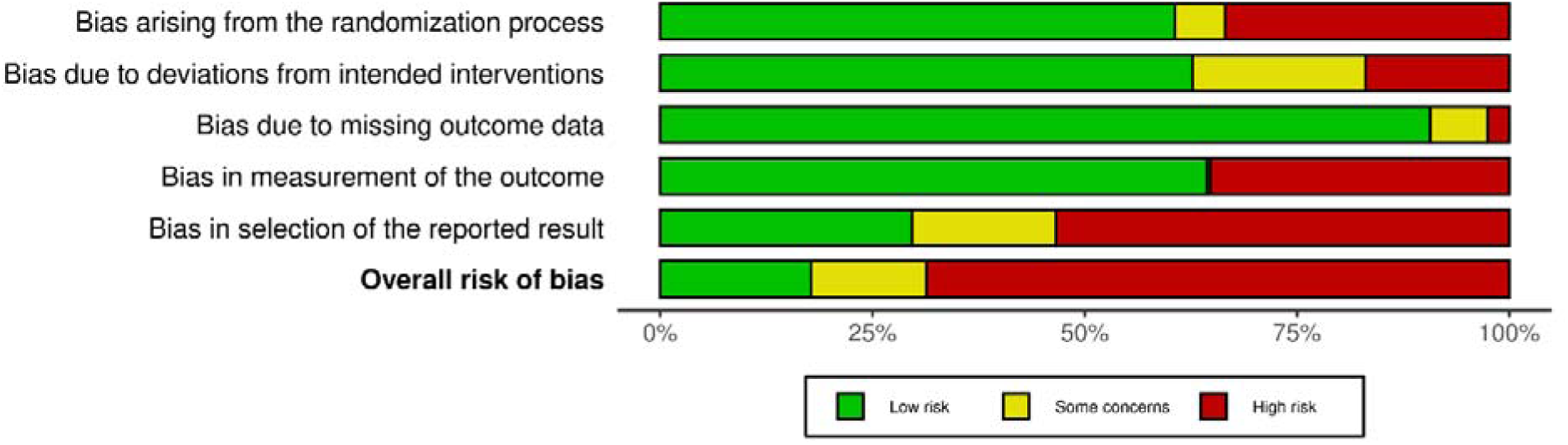
Summary of the Risk of bias with RoB-2 tool of the included RCTs.

In exploratory analysis, no association was found between conclusion of the abstract and overall RoB. Favorable conclusions were seen in 106 (69.7%) of the RCTs at high risk of bias, in 19 67.9%) of the RCTs with some concerns, and in 31 (75.6%) of the RCTs with low risk of bias (Supplementary Table 5). The corresponding number for unfavorable conclusions were 21 (13.8%), 4 (14.3%), and 4 (9.8%), respectively.

## Discussion

Our study found extensive heterogeneity in the features and characteristics of the interventions labeled as ‘personalized’, ‘individualized’, or, far less commonly, ‘precision’. Nevertheless, despite frequent conceptual distinctions in the literature, these labels were applied largely interchangeably across trials. Evaluation of these studies showed that most of them had some favorable results for the experimental intervention(s), very few had transparency in terms of data and code sharing, and most suffered from high risk of bias. Most interventions described with these terms relied on non-genomic features, such as lifestyle behaviors, psychological characteristics, disease classification, or functional capacity. Genetic and omics-based information has been often portrayed as the defining hallmark of precision medicine [23–25]. Precision oncology [26,27] has a strong focus on targeted pharmacological therapies based on single genetic mutations [28]. However, in our study, these applications were comparatively rare. This may reflect a substantial disconnect between dominant narratives in precision medicine discourse and the realities of clinical trial implementation. An alternative explanation is that precision oncology trials do not use that terminology in the titles of their articles and thus they were not included in our eligible RCTs. Regardless, the whole discipline of personalized/individualized/precision medicine is shifting towards an integrative approach that incorporates also lifestyle, environmental exposures, education, and socioeconomic factors [29–31], rather than just molecular profiling. In our sample of RCTs, interventions labeled as personalized, individualized, or precision were more frequently behavioral in nature and primarily based on lifestyle-related characteristics. These contextual elements are fundamental while personalizing interventions [32,33].

In our included interventions, personalization most often involved adjusting intervention content, intensity, or dosage based on readily observable or self-reported individual characteristics. Moreover, the predominance of single interventions tailored differently for each participant suggests that personalization in RCTs is frequently operationalized as flexible delivery rather than as fundamentally distinct therapeutic strategies [34]. In this sense, personalized interventions are often heterogeneous in their application but not in their underlying structure, design, or conceptual framework. More simply, the intervention remains essentially the same but is adapted to individual characteristics [34,35].

This approach is particularly relevant in studies involving rehabilitation [36] or physical exercise interventions [37]. Moving beyond rigid programs with identical targets for all participants may help overcome a common limitation of such interventions, namely the failure to achieve predefined goals [38]. In practice, objectives are often expressed as fixed thresholds, such as a minimum number of steps per day or minutes of physical activity. However, for individuals with different baseline characteristics, abilities, or health conditions, personalized targets may lead to greater adherence, improved health outcomes, or enhanced preventive effects, ultimately resulting in greater overall benefit [39,40]. While such tailoring may be clinically sensible, its methodological implications are important. When interventions vary extensively across participants, interpretability, reproducibility, and causal attribution become more challenging, particularly if tailoring algorithms are insufficiently described [41].

Among the published RCTs in our study, the conclusions of the Abstract almost always had some favorable elements (either exclusively favorable or mixed) for personalized interventions. Very few published trials reached only unfavorable conclusions. This extremely high rate of success seems spurious. It therefore raises concerns about possible biases that may have distorted, selected, and/or exaggerated the research findings of these studies. The threat of this literature being unreliable is increased given that the majority of studies were already judged to be at high risk of bias based on RoB2 assessments. Moreover, additional biases may exist beyond those captured by the RoB2 tool.

To safeguard this research literature and enhance trust in its findings, transparency would be most welcome. However, we also documented that critical transparency practices were also very limited, especially with regard to sharing of data and code. The very low rate of data sharing and the almost non-existent sharing of code places these trials at lower transparency ratings, compared with the average biomedical literature and other trials [18]. However, other studies of high-profile trials also show very low rates of data and code sharing (e.g. 4.4% and 4.5%, respectively in Siena et al. [42]). This lack of transparency is especially problematic in studies where intervention tailoring relies on decision rules, algorithms, or adaptive components that are central to the intervention itself [41]. Even though not all RCTs included in our review relied on complex algorithms, and in such cases code sharing may be less critical when the intervention is sufficiently described, enhanced sharing is currently feasible for RCTs in general [43] and should be promoted also for these specific trials.

Careful pre-registration with availability of detailed protocols with extensively documented statistical analysis plans would also enhance trust in this literature. In our eligible RCTs, we documented that three-quarters were registered, with less than 60% were registered before the trial started. While we did not examine in depth the registration entries, it is typical for registry entries of RCTs to give very limited information on details of the proposed analysis, and adherence to registration guidance is low [44–46]. Therefore, this poor documentation may leave many researcher degrees of freedom on what they report and what to conclude about the trial.

We observed no substantive differences in study characteristics based on whether authors used the term ‘personalized’, ‘individualized’, or ‘precision’. This supports the interpretation that these terms function largely as rhetorical labels [47,48], rather than as indicators of distinct methodological or practical approaches, and that in clinical research they are often used interchangeably [9]. In this respect, their use in practice diverges from theoretical definitions, particularly for personalized and precision medicine, which are frequently treated as distinct concepts in the literature and by experts [49,50].

### Limitations

This study has some limitations. First, our search was restricted to RCTs that included the relevant terms in the title, potentially excluding studies that implemented individualized approaches without explicitly labeling them as such. However, this choice is consistent with our objective of examining how these terms are publicly and prominently used in the scientific literature. Second, only a limited number of RCTs used the term ‘precision’, which restricts the generalizability of findings related to this label. Third, we focused on the three key terms denoting personalization that may apply to any field. Some additional terms are more specific to specific fields and interventions. It would be too convoluted to unearth every single trial that has personalization features. For example, some oncology trials with personalization features may use a variety of terms such as ‘targeted’ therapy, ‘master’ protocols, ‘umbrella’ and ‘basket’ designs, ‘enrichment’ or ‘adaptive’[51]. Finally, the 2020-2022 search period overlaps with the COVID-19 pandemic, which may have influenced the broader research landscape and publication trends, potentially affecting the representativeness of the included evidence.

### Conclusions

Contemporary RCTs that include interventions labeled as ‘personalized’, ‘individualized’, or ‘precision’ encompass a wide and heterogeneous set, and most rely on lifestyle and behavioral approaches. In practice, these terms are used largely interchangeably, reflecting a lack of clear distinction in their use in clinical research and practice. The fact that most of these trials reach favorable conclusions and have low transparency, and high risk of bias suggests that there is substantial room for improving the rigor and reliability of this important line of clinical and translational investigation. Our evaluated sample of 262 RCTs provides a recent baseline against which future progress in the field can be probed.

## Supporting information

Supplementary tables and List of included randomized controlled trials

## Declarations

## Acknowledgements

The authors would like to sincerely thank Dr. Salvatore Di Grande and Dr. Alessio Perilli for their support during the early phases of the study.

## Ethical approval

Not required (evaluation of published literature).

## Contributors

LR: Conceptualization, Methodology, Validation, Formal analysis, Investigation, Data Curation, Writing - Original Draft, Writing - Review & Editing, Visualization. NL: Validation, Formal analysis, Data Curation, Writing - Original Draft, Writing - Review & Editing, Visualization. LS: Formal analysis, Investigation, Writing - Review & Editing. RP: Conceptualization, Methodology, Writing - Original Draft, Writing - Review & Editing, Supervision. SB: Conceptualization, Methodology, Validation, Resources, Writing - Original Draft, Writing - Review & Editing, Supervision, funding acquisition. JPAI: Conceptualization, Methodology, Validation, Formal analysis, Writing - Original Draft, Writing - Review & Editing, Visualization, Supervision. JPAI and SB are the guarantors of the study.

## Data availability statement and code sharing

The full list of the included RCTs in contained in Supplementary File 1. The dataset used for the analysis will be available on Open Science Framework at this link: https://osf.io/cgy4k/overview.

## Transparency

The lead authors (the manuscript’s guarantors) affirm that the manuscript is an honest, accurate, and transparent account of the study being reported; that no important aspects of the study have been omitted; and that any discrepancies from the study as planned (and, if relevant, registered) have been explained.

