## Supplementary tables and List of included randomized controlled trials for "Randomized trials of ‘personalized’, ‘individualized’ and ‘precision’ interventions are very diverse and have low transparency and high bias"

**Supplementary Table S1**. Subgroup analysis for differences between favorable, unfavorable or mixed conclusions in the abstracts towards the personalized intervention.

| **Variable** | | N (%) | N (%) | N (%) | N (%) | |
|---|---|---|---|---|---|---|
| | | Overall | Favorable | Mixed | Unfavorable | p |
| **Included studies** | | 221 (100%) | 156 (70.6%) | 36 (16.3%) | 29 (13.1%) | - |
| **Year** | 2020 | 63 (28.5) | 47 (30.1) | 11 (30.6) | 5 (17.2) | 0.558 |
| | 2021 | 70 (31.7) | 51 (32.7) | 10 (27.8) | 9 (31.0) | |
| | 2022 | 88 (39.8) | 58 (37.2) | 15 (41.7) | 15 (51.7) | |
| **Country** | Europe | 68 (30.8%) | 53 (34.0%) | 6 (16.7%) | 9 (31.0%) | 0.023 |
| | UK | 8 (3.6) | 6 (3.8) | 1 (2.8) | 1 (3.4) | |
| | France | 13 (5.9) | 10 (6.4) | 3 (8.3) | 0 (0.0) | |
| | Germany | 10 (4.5) | 6 (3.8) | 0 (0.0) | 4 (13.8) | |
| | Europe Others | 37 (16.7) | 31 (19.9) | 2 (5.6) | 4 (13.8) | |
| | USA | 47 (21.3) | 31 (19.9) | 12 (33.3) | 4 (13.8) | |
| | China | 38 (17.2) | 31 (19.9) | 4 (11.1) | 3 (10.3) | |
| | Australia | 15 (6.8) | 6 (3.8) | 5 (13.9) | 4 (13.8) | |
| | Canada | 11 (5.0) | 6 (3.8) | 3 (8.3) | 2 (6.9) | |
| | World others | 42 (19.1) | 29 (18.6) | 6 (16.7) | 7 (24.1) | |
| **Medical specialty** | Cardiology | 27 (12.2) | 16 (10.3) | 6 (16.7) | 5 (17.2) | 0.012 |
| | Psychiatry | 25 (11.3) | 16 (10.3) | 3 (8.3) | 6 (20.7) | |
| | Endocrinology | 23 (10.4) | 16 (10.3) | 4 (11.1) | 3 (10.3) | |
| | Oncology | 23 (10.4) | 14 (9.0) | 7 (19.4) | 2 (6.9) | |
| | Obstetrics and gynecology | 15 (6.8) | 13 (8.3) | 2 (5.6) | 0 (0.0) | |
| | Orthopedics | 14 (6.3) | 8 (5.1) | 5 (13.9) | 1 (3.4) | |
| | Respiratory medicine | 13 (5.9) | 9 (5.8) | 2 (5.6) | 2 (6.9) | |
| | Infectious Diseases | 11 (5.0) | 6 (3.8) | 3 (8.3) | 2 (6.9) | |
| | Others | 74 (33.5%) | 58 (37.2%) | 4 (11.1%) | 8 (27.6%) | |
| **Number of arms** | 2 | 185 (83.7) | 129 (82.7) | 33 (91.7) | 23 (79.3) | 0.344 |
| | 3 | 27 (12.2) | 21 (13.5) | 2 (5.6) | 4 (13.8) | |
| | 4 | 8 (3.6) | 6 (3.8) | 1 (2.8) | 1 (3.4) | |
| | 6 | 1 (0.5) | 0 (0.0) | 0 (0.0) | 1 (3.4) | |
| **Total sample size** | median (IQR) | 135.00 [63.00, 300.00] | 120.00 [60.00, 270.75] | 168.50 [63.75, 302.50] | 197.00 [73.00, 484.00] | 0.165 |
| **Group of the study** | 1a | 202 (91.4%) | 141 (90.4%) | 34 (94.4%) | 27 (93.1%) | 0.866 |
| | 1b | 19 (8.6) | 15 (9.6) | 2 (5.6) | 2 (6.9) | |
| **Term used** | Individualized | 102 (46.2) | 71 (45.5) | 15 (41.7) | 16 (55.2) | 0.581 |
| | Personalized | 110 (49.8) | 79 (50.6) | 20 (55.6) | 11 (37.9) | |
| | Precision | 9 (4.1) | 6 (3.8) | 1 (2.8) | 2 (6.9) | |
| **Type of application** | Therapy | 153 (69.2) | 110 (70.5) | 23 (63.9) | 20 (69.0) | 0.822 |

| | | | | | | |
|---|---|---|---|---|---|---|
| | Secondary prevention | 66 (29.9) | 44 (28.2) | 13 (36.1) | 9 (31.0) | |
| | Primary prevention | 2 (0.9) | 2 (1.3) | 0 (0.0) | 0 (0.0) | |
| **Difference within the intervention arm [a]** | Dosage of therapy | 114 (51.6) | 83 (53.2) | 15 (41.7) | 16 (55.2) | 0.435 |
| | Schedule of therapy | 64 (29.0) | 43 (27.6) | 10 (27.8) | 11 (37.9) | 0.510 |
| | Drug used | 21 (9.5) | 16 (10.3) | 2 (5.6) | 3 (10.3) | 0.764 |
| | Testing | 17 (7.7) | 12 (7.7) | 3 (8.3) | 2 (6.9) | 1.000 |
| | Anatomical target | 11 (5.0) | 7 (4.5) | 3 (8.3) | 1 (3.4) | 0.613 |
| | Follow-up/visits | 11 (5.0) | 8 (5.1) | 1 (2.8) | 2 (6.9) | 0.785 |
| | Other | 43 (19.5) | 28 (17.9) | 9 (25.0) | 6 (20.7) | 0.574 |
| **Difference in dosage** | 1 | 4 (1.8) | 3 (1.9) | 0 (0.0) | 1 (3.4) | 0.404 |
| | 2 | 3 (1.4) | 3 (1.9) | 0 (0.0) | 0 (0.0) | |
| | 3 | 3 (1.4) | 3 (1.9) | 0 (0.0) | 0 (0.0) | |
| | 4 | 3 (1.4) | 3 (1.9) | 0 (0.0) | 0 (0.0) | |
| | >4 | 20 (9.0) | 9 (5.8) | 5 (13.9) | 6 (20.7) | |
| | For each patient | 91 (41.2) | 68 (43.6) | 12 (33.3) | 11 (37.9) | |
| | NA | 97 (43.9) | 67 (42.9) | 19 (52.8) | 11 (37.9) | |
| **Number of personalized intervention(s)** | 1 | 157 (71.0) | 110 (70.5) | 24 (66.7) | 23 (79.3) | 0.381 |
| | 2 | 26 (11.8) | 21 (13.5) | 4 (11.1) | 1 (3.4) | |
| | 3 | 12 (5.4) | 10 (6.4) | 2 (5.6) | 0 (0.0) | |
| | 4 | 2 (0.9) | 1 (0.6) | 1 (2.8) | 0 (0.0) | |
| | >4 | 24 (10.9) | 14 (9.0) | 5 (13.9) | 5 (17.2) | |
| **Difference in tailoring** | 1 | 1 (0.5) | 1 (0.6) | 0 (0.0) | 0 (0.0) | 0.355 |
| | 2 | 4 (1.8) | 4 (2.6) | 0 (0.0) | 0 (0.0) | |
| | 3 | 2 (0.9) | 2 (1.3) | 0 (0.0) | 0 (0.0) | |
| | 4 | 1 (0.5) | 1 (0.6) | 0 (0.0) | 0 (0.0) | |
| | >4 | 18 (8.1) | 8 (5.1) | 5 (13.9) | 5 (17.2) | |
| | For each patient | 182 (82.4) | 129 (82.7) | 31 (86.1) | 22 (75.9) | |
| | NA | 13 (5.9) | 11 (7.1) | 0 (0.0) | 2 (6.9) | |
| **Type of intervention [a]** | Behavioral | 75 (33.9%) | 52 (33.3%) | 15 (41.7%) | 8 (27.6%) | 0.486 |
| | Digital | 65 (29.4%) | 46 (29.5%) | 9 (25.0%) | 10 (34.5%) | 0.695 |
| | Medication | 60 (27.1%) | 37 (23.7%) | 9 (25.0%) | 14 (48.3%) | 0.028 |
| | Psychological | 47 (21.3%) | 32 (20.5%) | 8 (22.2%) | 7 (24.1%) | 0.872 |
| | Physical exercises | 34 (15.4%) | 28 (17.9%) | 5 (13.9%) | 1 (3.4%) | 0.125 |
| | Dietary | 30 (13.6%) | 26 (16.7%) | 2 (5.6%) | 2 (6.9%) | 0.131 |
| | Educational | 31 (14.0%) | 24 (15.4%) | 4 (11.1%) | 3 (10.3%) | 0.716 |
| | Rehabilitation | 19 (8.6%) | 16 (10.3%) | 2 (5.6%) | 1 (3.4%) | 0.556 |
| | Surgery | 15 (6.8%) | 11 (7.1%) | 1 (2.8%) | 3 (10.3%) | 0.485 |
| | Anesthesia | 15 (6.8%) | 11 (7.1%) | 2 (5.6%) | 2 (6.9%) | 1.000 |
| | Omics | 6 (2.7%) | 4 (2.6%) | 2 (5.6%) | 0 (0.0%) | 0.405 |
| | Physiotherapy | 4 (1.8%) | 2 (1.3%) | 1 (2.8%) | 1 (3.4%) | 0.337 |

| | | | | | | |
|---|---|---|---|---|---|---|
| | Radiotherapy | 3 (1.4%) | 2 (1.3%) | 1 (2.8%) | 0 (0.0%) | 0.650 |
| **Main category of individual features** [a] | Lifestyle and behavioral | 85 (38.5) | 61 (39.1) | 19 (52.8) | 5 (17.2) | 0.013 |
| | Disease classification | 61 (27.6) | 45 (28.8) | 11 (30.6) | 5 (17.2) | 0.420 |
| | Psychological | 58 (26.2) | 40 (25.6) | 11 (30.6) | 7 (24.1) | 0.813 |
| | Vital signs | 49 (22.2) | 38 (24.4) | 5 (13.9) | 6 (20.7) | 0.433 |
| | Physical or functional capacity | 47 (21.3) | 36 (23.1) | 7 (19.4) | 4 (13.8) | 0.597 |
| | Anthropometrics | 36 (16.3) | 28 (17.9) | 4 (11.1) | 4 (13.8) | 0.656 |
| | Test on biological material | 34 (15.4) | 26 (16.7) | 2 (5.6) | 6 (20.7) | 0.165 |
| | Imaging tools | 23 (10.4) | 18 (11.5) | 2 (5.6) | 3 (10.3) | 0.610 |
| | Demographics | 22 (10.0) | 16 (10.3) | 4 (11.1) | 2 (6.9) | 0.882 |
| | Complex scores (specify) | 20 (9.0) | 12 (7.7) | 4 (11.1) | 4 (13.8) | 0.431 |
| | Genetic | 13 (5.9) | 5 (3.2) | 7 (19.4) | 1 (3.4) | **0.003** |
| | Socioeconomic | 11 (5.0) | 6 (3.8) | 4 (11.1) | 1 (3.4) | 0.182 |
| | Omics | 8 (3.6) | 5 (3.2) | 3 (8.3) | 0 (0.0) | 0.194 |
| | Other | 16 (7.2) | 11 (7.1) | 2 (5.6) | 3 (10.3) | 0.713 |
| **subcategories** [a] | Patient preferences | 34 (15.4) | 24 (15.4) | 7 (19.4) | 3 (10.3) | 0.614 |
| | Weight | 31 (14.0) | 24 (15.4) | 3 (8.3) | 4 (13.8) | 0.648 |
| | Blood | 27 (12.2) | 21 (13.5) | 2 (5.6) | 4 (13.8) | 0.441 |
| | Motivation | 18 (8.1) | 12 (7.7) | 4 (11.1) | 2 (6.9) | 0.739 |
| | Blood pressure | 14 (6.3) | 12 (7.7) | 2 (5.6) | 0 (0.0) | 0.377 |
| | Education | 11 (5.0) | 6 (3.8) | 4 (11.1) | 1 (3.4) | 0.182 |
| | Pulse rate | 11 (5.0) | 10 (6.4) | 0 (0.0) | 1 (3.4) | 0.300 |
| | Respiration rate | 8 (3.6) | 6 (3.8) | 1 (2.8) | 1 (3.4) | 1.000 |
| | Genetic testing | 6 (2.7) | 3 (1.9) | 3 (8.3) | 0 (0.0) | 0.108 |
| | BMI | 5 (2.3) | 4 (2.6) | 1 (2.8) | 0 (0.0) | 1.000 |
| | Lifetime risk | 4 (1.8) | 2 (1.3) | 2 (5.6) | 0 (0.0) | 0.207 |
| | Age | 2 (0.9) | 0 (0.0) | 1 (2.8) | 1 (3.4) | 0.086 |
| | Biopsy | 2 (0.9) | 1 (0.6) | 0 (0.0) | 1 (3.4) | 0.272 |
| | Metabolomics | 2 (0.9) | 2 (1.3) | 0 (0.0) | 0 (0.0) | 1.000 |
| | Temperature | 2 (0.9) | 1 (0.6) | 0 (0.0) | 1 (3.4) | 0.272 |
| | Genetic risk score (GRS) | 1 (0.5) | 0 (0.0) | 1 (2.8) | 0 (0.0) | 0.294 |
| | Microbiome | 1 (0.5) | 1 (0.6) | 0 (0.0) | 0 (0.0) | 1.000 |
| | Nutrigenomics | 1 (0.5) | 1 (0.6) | 0 (0.0) | 0 (0.0) | 1.000 |
| | Proteomics | 1 (0.5) | 1 (0.6) | 0 (0.0) | 0 (0.0) | 1.000 |
| | Urine | 1 (0.5) | 1 (0.6) | 0 (0.0) | 0 (0.0) | 1.000 |
| | Other | 2 (1.3) | 0 (0.0) | 0 (0.0) | 2 (0.9) | 0.017 |

[a] = Same studies employed multiple measurements. NA=Not Applicable. In bold p<0.005.

**Supplementary Table S2**. Subgroup analysis for differences between study design (trial vs protocol).

| **Variable** | | N (%) | N (%) | N (%) | |
|---|---|---|---|---|---|
| | | Overall | Protocol | Trial | p |
| **Included studies** | | 262 | 27 | 235 | - |
| **Year** | 2020 | 78 (29.8) | 11 (40.7) | 67 (28.5) | 0.441 |
| | 2021 | 82 (31.3) | 7 (25.9) | 75 (31.9) | |
| | 2022 | 102 (38.9) | 9 (33.3) | 93 (39.6) | |
| **Country** | Europe | 86 (32.8%) | 14 (51.9%) | 72 (30.6%) | 0.130 |
| | Germany | 14 (5.3) | 5 (18.5) | 9 (3.8) | |
| | UK | 13 (5.0) | 0 (0.0) | 13 (5.5) | |
| | France | 12 (4.6) | 1 (3.7) | 11 (4.7) | |
| | Europe Others | 47 (17.9) | 8 (29.6) | 39 (16.6) | |
| | USA | 58 (22.1) | 5 (18.5) | 53 (22.6) | |
| | China | 42 (16.0) | 3 (11.1) | 39 (16.6) | |
| | Australia | 19 (7.3) | 3 (11.1) | 16 (6.8) | |
| | Canada | 12 (4.6) | 1 (3.7) | 11 (4.7) | |
| | World others | 45 (17.2) | 1 (3.7) | 44 (18.7) | |
| **Medical specialty** | Cardiology | 32 (12.2) | 5 (18.5) | 27 (11.5) | 0.513 |
| | Psychiatry | 29 (11.1) | 2 (7.4) | 27 (11.5) | |
| | Oncology | 28 (10.7) | 5 (18.5) | 23 (9.8) | |
| | Endocrinology | 27 (10.3) | 4 (14.8) | 23 (9.8) | |
| | Obstetrics and gynecology | 16 (6.1) | 0 (0.0) | 16 (6.8) | |
| | Orthopedics | 16 (6.1) | 1 (3.7) | 15 (6.4) | |
| | Infectious Diseases | 14 (5.3) | 2 (7.4) | 12 (5.1) | |
| | Internal medicine | 13 (5.0) | 2 (7.4) | 11 (4.7) | |
| | Respiratory medicine | 13 (5.0) | 0 (0.0) | 13 (5.5) | |
| | Others | 72 (27.5%) | 6 (22.2%) | 66 (28.1%) | |
| **Number of arms** | 2 | 221 (84.4) | 24 (88.9) | 197 (83.8) | 1.000 |
| | 3 | 32 (12.2) | 3 (11.1) | 29 (12.3) | |
| | 4 | 8 (3.1) | 0 (0.0) | 8 (3.4) | |
| | 6 | 1 (0.4) | 0 (0.0) | 1 (0.4) | |
| **Total sample size** | median (IQR) | 136.00 [63.25, 302.25] | 242.00 [100.00, 383.00] | 128.00 [63.00, 293.50] | 0.081 |
| **Group of the study** | 1a | 225 (85.9) | 23 (85.2) | 202 (86.0) | 0.905 |
| | 1b | 21 (8.0) | 2 (7.4) | 19 (8.1) | |
| | 2 | 16 (6.1) | 2 (7.4) | 14 (6.0) | |
| **Term used** | Individualized | 120 (45.8) | 10 (37.0) | 110 (46.8) | 0.048 |
| | Personalized | 129 (49.2) | 13 (48.1) | 116 (49.4) | |
| | Precision | 13 (5.0) | 4 (14.8) | 9 (3.8) | |

| | | | | | |
|---|---|---|---|---|---|
| **Type of application** | Therapy | 186 (71.0) | 24 (88.9) | 162 (68.9) | 0.05 |
| | Secondary prevention | 74 (28.2) | 3 (11.1) | 71 (30.2) | |
| | Primary prevention | 2 (0.8) | 0 (0.0) | 2 (0.9) | |
| **Difference within the intervention arm [a]** | Dosage of therapy | 136 (40.5) | 14 (38.9) | 122 (40.7) | 1.000 |
| | Schedule of therapy | 75 (22.3) | 9 (25.0) | 66 (22.0) | 0.653 |
| | Drug used | 32 (9.5) | 8 (22.2) | 24 (8.0) | 0.009 |
| | Testing | 18 (5.4) | 1 (2.8) | 17 (5.7) | 0.704 |
| | Anatomical target | 14 (4.2) | 1 (2.8) | 13 (4.3) | 1.000 |
| | Follow-up/visits | 14 (4.2) | 2 (5.6) | 12 (4.0) | 0.644 |
| | Other | 47 (14.0) | 1 (2.8) | 46 (15.3) | 0.059 |
| **Difference in dosage** | 1 | 4 (1.5) | 0 (0.0) | 4 (1.7) | 0.594 |
| | 2 | 4 (1.5) | 1 (3.7) | 3 (1.3) | |
| | 3 | 3 (1.1) | 0 (0.0) | 3 (1.3) | |
| | 4 | 4 (1.5) | 1 (3.7) | 3 (1.3) | |
| | >4 | 25 (9.5) | 2 (7.4) | 23 (9.8) | |
| | For each patient | 109 (41.6) | 13 (48.1) | 96 (40.9) | |
| | NA | 113 (43.1) | 10 (37.0) | 103 (43.8) | |
| **Number of personalized intervention(s)** | 1 | 177 (67.6) | 18 (66.7) | 159 (67.7) | 0.352 |
| | 2 | 42 (16.0) | 6 (22.2) | 36 (15.3) | |
| | 3 | 13 (5.0) | 0 (0.0) | 13 (5.5) | |
| | 4 | 3 (1.1) | 1 (3.7) | 2 (0.9) | |
| | >4 | 27 (10.3) | 2 (7.4) | 25 (10.6) | |
| **Difference in tailoring** | 1 | 1 (0.4) | 0 (0.0) | 1 (0.4) | 1.000 |
| | 2 | 5 (1.9) | 0 (0.0) | 5 (2.1) | |
| | 3 | 2 (0.8) | 0 (0.0) | 2 (0.9) | |
| | 4 | 1 (0.4) | 0 (0.0) | 1 (0.4) | |
| | >4 | 21 (8.0) | 2 (7.4) | 19 (8.1) | |
| | For each patient | 217 (82.8) | 24 (88.9) | 193 (82.1) | |
| | NA | 15 (5.7) | 1 (3.7) | 14 (6.0) | |
| **Type of intervention [a]** | Behavioral | 87 (33.2%) | 8 (29.6%) | 79 (33.6%) | 0.830 |
| | Digital | 77 (29.4%) | 10 (37.0%) | 67 (28.5%) | 0.376 |
| | Medication | 72 (27.5%) | 9 (33.3%) | 63 (26.8%) | 0.498 |
| | Psychological | 57 (21.8%) | 7 (25.9%) | 50 (21.3%) | 0.623 |
| | Physical exercises | 44 (16.8%) | 7 (25.9%) | 37 (15.7%) | 0.181 |
| | Dietary | 41 (15.6%) | 7 (25.9%) | 34 (14.5%) | 0.157 |
| | Educational | 33 (12.6%) | 1 (3.7%) | 32 (13.6%) | 0.219 |
| | Rehabilitation | 22 (8.4%) | 1 (3.7%) | 21 (8.9%) | 0.711 |
| | Surgery | 17 (6.5%) | 1 (3.7%) | 16 (6.8%) | 1.000 |
| | Anesthesia | 16 (6.1%) | 0 (0.0%) | 16 (6.8%) | 0.386 |
| | Omics | 6 (2.3%) | 0 (0.0%) | 6 (2.6%) | 1.000 |

| | | | | | |
|---|---|---|---|---|---|
| | Physiotherapy | 4 (1.5%) | 0 (0.0%) | 4 (1.7%) | 1.000 |
| | Radiotherapy | 4 (1.5%) | 1 (3.7%) | 3 (1.3%) | 0.354 |
| **Main category of individual features** [a] | Lifestyle and behavioral | 98 (37.4) | 7 (25.9) | 91 (38.7) | 0.215 |
| | Disease classification | 75 (28.6) | 11 (40.7) | 64 (27.2) | 0.176 |
| | Psychological | 71 (27.1) | 10 (37.0) | 61 (26.0) | 0.254 |
| | Vital signs | 56 (21.4) | 5 (18.5) | 51 (21.7) | 0.809 |
| | Physical or functional capacity | 55 (21.0) | 6 (22.2) | 49 (20.9) | 0.807 |
| | Anthropometrics | 47 (17.9) | 8 (29.6) | 39 (16.6) | 0.112 |
| | Test on biological material | 42 (16.0) | 6 (22.2) | 36 (15.3) | 0.404 |
| | Imaging tools | 29 (11.1) | 4 (14.8) | 25 (10.6) | 0.516 |
| | Demographics | 24 (9.2) | 1 (3.7) | 23 (9.8) | 0.485 |
| | Complex scores (specify) | 22 (8.4) | 2 (7.4) | 20 (8.5) | 1.000 |
| | Genetic | 14 (5.3) | 2 (7.4) | 12 (5.1) | 0.644 |
| | Socioeconomic | 13 (5.0) | 0 (0.0) | 13 (5.5) | 0.373 |
| | Omics | 8 (3.1) | 0 (0.0) | 8 (3.4) | 1.000 |
| | Other | 2 (7.4) | 17 (7.2) | 17 (7.4%) | 0.000 |
| **Subcategories** [a] | Patient preferences | 41 (15.6) | 6 (22.2) | 35 (14.9) | 0.398 |
| | Weight | 41 (15.6) | 8 (29.6) | 33 (14.0) | 0.048 |
| | Blood | 35 (13.4) | 6 (22.2) | 29 (12.3) | 0.226 |
| | Motivation | 25 (9.5) | 6 (22.2) | 19 (8.1) | 0.030 |
| | Blood pressure | 17 (6.5) | 2 (7.4) | 15 (6.4) | 0.690 |
| | Education | 14 (5.3) | 2 (7.4) | 12 (5.1) | 0.644 |
| | Pulse rate | 12 (4.6) | 1 (3.7) | 11 (4.7) | 1.000 |
| | Respiration rate | 8 (3.1) | 0 (0.0) | 8 (3.4) | 1.000 |
| | Genetic testing | 7 (2.7) | 2 (7.4) | 5 (2.1) | 0.155 |
| | BMI | 6 (2.3) | 0 (0.0) | 6 (2.6) | 1.000 |
| | Lifetime risk | 5 (1.9) | 1 (3.7) | 4 (1.7) | 0.422 |
| | Age | 3 (1.1) | 1 (3.7) | 2 (0.9) | 0.279 |
| | Biopsy | 2 (0.8) | 0 (0.0) | 2 (0.9) | 1.000 |
| | Metabolomics | 2 (0.8) | 0 (0.0) | 2 (0.9) | 1.000 |
| | Temperature | 2 (0.8) | 0 (0.0) | 2 (0.9) | 1.000 |
| | Genetic Risk Score (GRS) | 1 (0.4) | 0 (0.0) | 1 (0.4) | 1.000 |
| | Microbiome | 1 (0.4) | 0 (0.0) | 1 (0.4) | 1.000 |
| | Nutrigenomics | 1 (0.4) | 0 (0.0) | 1 (0.4) | 1.000 |
| | Proteomics | 1 (0.4) | 0 (0.0) | 1 (0.4) | 1.000 |
| | Urine | 1 (0.4) | 0 (0.0) | 1 (0.4) | 1.000 |
| | Other | 2 (0.8) | 0 (0.0) | 2 (0.9) | 1.000 |

[a]= Same studies employed multiple measurements. NA=Not Applicable. In bold $p<0.005$.

**Supplementary Table 3.** Association analysis between features and categories

| Variable | | Term used | Abstract conclusions | Study design |
|---|---|---|---|---|
| **Year** | - | 0.517 | 0.558 | 0.441 |
| **Country** | - | <0.001 | 0.023 | 0.130 |
| **Medical specialty** | - | 0.017 | 0.012 | 0.513 |
| **Number of trial arms** | - | 0.787 | 0.344 | 1.000 |
| **Sample size** | - | 0.045 | 0.165 | 0.081 |
| **Study group** | - | 0.138 | 0.866 | 0.905 |
| **Type of application** | - | 0.069 | 0.822 | 0.05 |
| **Difference within the intervention arm** [a] | Dosage of therapy | 0.055 | 0.435 | 1.000 |
| | Schedule of therapy | 0.027 | 0.510 | 0.653 |
| | drug used | 0.069 | 0.764 | 0.009 |
| | testing | 0.064 | 1.000 | 0.704 |
| | Anatomical target | 0.897 | 0.613 | 1.000 |
| | Follow-up/visits | 0.427 | 0.785 | 0.644 |
| | Other | 0.188 | 0.574 | 0.059 |
| **Difference in dosage** | - | 0.015 | 0.404 | 0.594 |
| **Number of personalized interventions** | - | 0.015 | 0.381 | 0.352 |
| **Difference in tailoring** | - | 0.014 | 0.355 | 1.000 |
| **Type of intervention** [a] | behavioral | **0.002** | 0.486 | 0.830 |
| | digital | **0.000** | 0.695 | 0.376 |
| | medication | **0.002** | 0.028 | 0.498 |
| | psychological | 0.024 | 0.872 | 0.623 |
| | physical exercises | 0.161 | 0.125 | 0.181 |
| | dietary | 0.181 | 0.131 | 0.157 |
| | educational | 0.659 | 0.716 | 0.219 |
| | rehabilitation | 0.514 | 0.556 | 0.711 |
| | surgery | 0.132 | 0.485 | 1.000 |
| | anesthesia | 0.015 | 1.000 | 0.386 |
| | omics | 0.034 | 0.405 | 1.000 |
| | physiotherapy | 0.474 | 0.337 | 1.000 |
| | radiotherapy | 0.474 | 0.650 | 0.354 |
| **Main category of individual features** [a] | Lifestyle and behavioral | 0.125 | 0.164 | 0.325 |
| | Psychological | 0.175 | 0.632 | 0.406 |
| | Disease classification | 0.711 | 0.414 | 0.292 |
| | Vital signs | 0.096 | 0.530 | 0.812 |
| | Physical or functional capacity | 0.187 | 0.665 | 0.812 |
| | Anthropometrics | 0.009 | 0.585 | 0.330 |

| | | | | |
|---|---|---|---|---|
| | Test on biological material | 0.894 | 0.206 | 0.315 |
| | Imaging tools | 0.168 | 0.764 | 0.522 |
| | Complex scores | 0.590 | 0.565 | 1.000 |
| | Demographics | 0.211 | 0.712 | 0.487 |
| | Other | 0.520 | 0.703 | 1.000 |
| | Genetic | 0.003 | 0.022 | 0.378 |
| | Socioeconomic | 0.031 | 0.212 | 0.658 |
| | Omics | 0.085 | 0.341 | 0.604 |
| **Subcategories [a]** | Weight | **0.002** | 0.679 | 0.679 |
| | Patient preferences | 0.736 | 0.755 | 0.755 |
| | Blood | 1.000 | 0.418 | 0.418 |
| | Motivation | 0.405 | 0.507 | 0.507 |
| | Blood pressure | 0.238 | 0.462 | 0.462 |
| | Pulse rate | 0.555 | 0.341 | 0.341 |
| | Education | 0.031 | 0.212 | 0.212 |
| | Respiration rate | 0.193 | 0.283 | 0.283 |
| | Genetic testing | 0.009 | 1.000 | 1.000 |
| | Lifetime risk | 0.053 | 0.298 | 0.298 |
| | BMI | 1.000 | 1.000 | 1.000 |
| | Metabolomics | 1.000 | 1.000 | 1.000 |
| | Age | 1.000 | 0.084 | 0.084 |
| | Biopsy | 0.552 | 0.257 | 0.257 |
| | Temperature | 1.000 | 0.257 | 0.257 |
| | Proteomics | 0.112 | 1.000 | 1.000 |
| | Nutrigenomics | 1.000 | 1.000 | 1.000 |
| | Microbiome | 1.000 | 1.000 | 1.000 |
| | Urine | 1.000 | 1.000 | 1.000 |
| | Genetic Risk Score (GRS) | 1.000 | 0.288 | 0.288 |
| | Other | 0.314 | 1.000 | 1.000 |

[a]= Same studies employed multiple measurements.

**Supplementary Table 4**. Summary of the Risk of bias with RoB-2 tool of the included RCTs.

| | D1 (N (%)) | D2 (N (%)) | D3 (N (%)) | D4 (N (%)) | D5 (N (%)) | Overall (N (%)) |
|---|---|---|---|---|---|---|
| Low | 143 (60.6) | 148 (62.7) | 214 (90.7) | 152 (64.4) | 70 (29.7) | 42 (17.8) |
| Some concerns | 14 (5.9) | 48 (20.3) | 16 (6.8) | 1 (0.4) | 40 (16.9) | 32 (13.6) |
| High | 79 (33.5) | 40 (16.9) | 6 (2.5) | 83 (35.2) | 126 (53.4) | 162 (68.6) |

**Supplementary Table 5. E**xploratory analysis of association between conclusion of the abstract and overall RoB.

| | **Risk of Bias** | | | **p-value** |
|---|---|---|---|---|
| **Conclusions** | High (N (%)) | Some concerns (N (%)) | Low (N (%)) | |
| Favorable for the personalized intervention | 106 (69.7%) | 19 (67.9%) | 31 (75.6%) | 0.489 |
| Mixed conclusions | 25 (16.4%) | 5 (17.9%) | 6 (14.6%) | |
| Unfavorable for the personalized intervention | 21 (13.8%) | 4 (14.3%) | 4 (9.8%) | |

**Supplementary file 1.** List of included RCTs

1. Leventogiannis. Toward personalized immunotherapy in sepsis: The PROVIDE randomized clinical trial. Cell reports. Medicine. 2022. 10.1016/j.xcrm.2022.100817.
2. Shah. Supervised, individualised exercise reduces fatigue and improves strength and quality of life more than unsupervised home exercise in people with chronic Guillain-Barre syndrome: a randomised trial. Journal of physiotherapy. 2022. 10.1016/j.jphys.2022.03.007.
3. Garin. Personalised versus standard dosimetry approach of selective internal radiation therapy in patients with locally advanced hepatocellular carcinoma (DOSISPHERE-01): a randomised, multicentre, open-label phase 2 trial. The lancet. Gastroenterology & hepatology. 2021. 10.1016/S2468-1253(20)30290-9.
4. Li. Personalized Variable vs Fixed-Dose Systemic Corticosteroid Therapy in Hospitalized Patients With Acute Exacerbations of COPD: A Prospective, Multicenter, Randomized, Open-Label Clinical Trial. Chest. 2021. 10.1016/j.chest.2021.05.024.
5. Bohé. Individualised versus conventional glucose control in critically-ill patients: the CONTROLING study-a randomized clinical trial. Intensive care medicine. 2021. 10.1007/s00134-021-06526-8.
6. Popp. Effect of a Personalized Diet to Reduce Postprandial Glycemic Response vs a Low-fat Diet on Weight Loss in Adults With Abnormal Glucose Metabolism and Obesity: A Randomized Clinical Trial. JAMA network open. 2022. 10.1001/jamanetworkopen.2022.33760.
7. Wu. Niraparib maintenance therapy in patients with platinum-sensitive recurrent ovarian cancer using an individualized starting dose (NORA): a randomized, double-blind, placebo-controlled phase III trial(â˜†). Annals of oncology : official journal of the European Society for Medical Oncology. 2021. 10.1016/j.annonc.2020.12.018.
8. Hersberger. Individualized Nutritional Support for Hospitalized Patients With Chronic Heart Failure. Journal of the American College of Cardiology. 2021. 10.1016/j.jacc.2021.03.232.
9. Garg. Personalised cooler dialysate for patients receiving maintenance haemodialysis (MyTEMP): a pragmatic, cluster-randomised trial. Lancet (London, England). 2022. 10.1016/S0140-6736(22)01805-0.
10. Ben-Yacov. Personalized Postprandial Glucose Response-Targeting Diet Versus Mediterranean Diet for Glycemic Control in Prediabetes. Diabetes care. 2021. 10.2337/dc21-0162.
11. Simón. A 5-year multicentre randomized controlled trial comparing personalized, frozen and fresh blastocyst transfer in IVF. Reproductive biomedicine online. 2020. 10.1016/j.rbmo.2020.06.002.
12. Rein. Effects of personalized diets by prediction of glycemic responses on glycemic control and metabolic health in newly diagnosed T2DM: a randomized dietary intervention pilot trial. BMC medicine. 2022. 10.1186/s12916-022-02254-y.
13. Roggeveen. Right dose, right now: bedside, real-time, data-driven, and personalised antibiotic dosing in critically ill patients with sepsis or septic shock-a two-centre randomised clinical trial. Critical care (London, England). 2022. 10.1186/s13054-022-04098-7.
14. Ewoldt. Model-informed precision dosing of beta-lactam antibiotics and ciprofloxacin in critically ill patients: a multicentre randomised clinical trial. Intensive care medicine. 2022. 10.1007/s00134-022-06921-9.
15. Lee. Efficacy of Personalized Diabetes Self-care Using an Electronic Medical Record-Integrated Mobile App in Patients With Type 2 Diabetes: 6-Month Randomized Controlled Trial. Journal of medical Internet research. 2022. 10.2196/37430.
16. McCreedy. Measuring the effects of a personalized music intervention on agitated behaviors among nursing home residents with dementia: design features for cluster-randomized adaptive trial. Trials. 2021. 10.1186/s13063-021-05620-y.
17. Hiensch. Design of a multinational randomized controlled trial to assess the effects of structured and individualized exercise in patients with metastatic breast cancer on fatigue and quality of life: the EFFECT study. Trials. 2022. 10.1186/s13063-022-06556-7.

18. Qiao. A randomised controlled trial to clinically validate follitropin delta in its individualised dosing regimen for ovarian stimulation in Asian IVF/ICSI patients. Human reproduction (Oxford, England). 2021. 10.1093/humrep/deab155.
19. Rodriguez. Community health workers and precision medicine: A randomized controlled trial Contemporary clinical trials. 2022. 10.1016/j.cct.2022.106906.
20. Joosten. Computer-assisted Individualized Hemodynamic Management Reduces Intraoperative Hypotension in Intermediate- and High-risk Surgery: A Randomized Controlled Trial. Anesthesiology. 2021. 10.1097/ALN.0000000000003807.
21. Bech. The effect of group or individualised pelvic floor exercises with or without ultrasonography guidance for urinary incontinence in elderly women - A pilot study. Journal of bodywork and movement therapies. 2021. 10.1016/j.jbmt.2021.07.032.
22. Calderon-Casellas. Assessment of skin cancer precision prevention materials among Hispanics in Florida and Puerto Rico. Patient education and counseling. 2022. 10.1016/j.pec.2022.06.012.
23. Fischer. Individualised or liberal red blood cell transfusion after cardiac surgery: a randomised controlled trial. British journal of anaesthesia. 2022. 10.1016/j.bja.2021.09.037.
24. Huo. Evaluation of individualized treatment of nonproliferative diabetic retinopathy: a multicenter, randomized, parallel-controlled study. Journal of traditional Chinese medicine = Chung i tsa chih ying wen pan. 2022. 10.19852/j.cnki.jtcm.20210425.002.
25. Liew. Understanding how individualised physiotherapy or advice altered different elements of disability for people with low back pain using network analysis. PloS one. 2022. 10.1371/journal.pone.0263574.
26. Bolognese. Group Nutrition Counseling or Individualized Prescription for Women With Obesity? A Clinical Trial. Frontiers in public health. 2020. 10.3389/fpubh.2020.00127.
27. Ma. Effect of individualized treatment strategy on postoperative nausea and vomiting in gynaecological laparoscopic surgery: a double-blind, randomized, controlled trial. BMC anesthesiology. 2022. 10.1186/s12871-022-01809-z.
28. Broadbent. Safety and cost-effectiveness of individualised screening for diabetic retinopathy: the ISDR open-label, equivalence RCT. Diabetologia. 2021. 10.1007/s00125-020-05313-2.
29. Teo. Efficacy of a novel personalised aflibercept monotherapy regimen based on polypoidal lesion closure in participants with polypoidal choroidal vasculopathy. The British journal of ophthalmology. 2022. 10.1136/bjophthalmol-2020-318354.
30. Lacson. A randomized clinical trial of precision prevention materials incorporating MC1R genetic risk to improve skin cancer prevention activities among Hispanics. Cancer research communications. 2022. 10.1158/2767-9764.crc-21-0114.
31. Beleigoli. Personalized Web-Based Weight Loss Behavior Change Program With and Without Dietitian Online Coaching for Adults With Overweight and Obesity: Randomized Controlled Trial. Journal of medical Internet research. 2020. 10.2196/17494.
32. Heeger. Individualized or fixed approach to pulmonary vein isolation utilizing the fourth-generation cryoballoon in patients with paroxysmal atrial fibrillation: the randomized INDI-FREEZE trial. Europace : European pacing, arrhythmias, and cardiac electrophysiology : journal of the working groups on cardiac pacing, arrhythmias, and cardiac cellular electrophysiology of the European Society of Cardiology. 2022. 10.1093/europace/euab305.
33. Xu. Effect of individualized weight management intervention on excessive gestational weight gain and perinatal outcomes: a randomized controlled trial. PeerJ. 2022. 10.7717/peerj.13067.
34. McCreedy. Pragmatic Trial of Personalized Music for Agitation and Antipsychotic Use in Nursing Home Residents With Dementia. Journal of the American Medical Directors Association. 2022. 10.1016/j.jamda.2021.12.030.
35. López. Effectiveness of an individualized comprehensive rehabilitation program in women with chronic knee osteoarthritis: a randomized controlled trial. Menopause (New York, N.Y.). 2022. 10.1097/GME.0000000000001959.

36. Zhang. Driving Pressure-Guided Individualized Positive End-Expiratory Pressure in Abdominal Surgery: A Randomized Controlled Trial. Anesthesia and analgesia. 2021. 10.1213/ANE.0000000000005575.
37. Omura. Effects of individualized dietary advice compared with conventional dietary advice for adults with type 2 diabetes: A randomized controlled trial. Nutrition, metabolism, and cardiovascular diseases : NMCD. 2022. 10.1016/j.numecd.2021.11.006.
38. Bertacco. Effect of personalized musical intervention on burden of care in dental implant surgery: A pilot randomized controlled trial. Journal of dentistry. 2022. 10.1016/j.jdent.2022.104091.
39. Nicklas. Personalised haemodynamic management targeting baseline cardiac index in high-risk patients undergoing major abdominal surgery: a randomised single-centre clinical trial. British journal of anaesthesia. 2020. 10.1016/j.bja.2020.04.094.
40. Ko. Personalized Behavioral Nutrition Among Older Asian Americans: Study Protocol. Nursing research. 2021. 10.1097/NNR.0000000000000514.
41. Martini. Changes in the Second Ventilatory Threshold Following Individualised versus Standardised Exercise Prescription among Physically Inactive Adults: A Randomised Trial. International journal of environmental research and public health. 2022. 10.3390/ijerph19073962.
42. Wernli. Effect of Personalized Breast Cancer Risk Tool on Chemoprevention and Breast Imaging: ENGAGED-2 Trial. JNCI cancer spectrum. 2021. 10.1093/jncics/pkaa114.
43. Jaspers. Communicating personalised statin therapy-effects as 10-year CVD-risk or CVD-free life-expectancy: does it improve decisional conflict? Three-armed, blinded, randomised controlled trial. BMJ open. 2021. 10.1136/bmjopen-2020-041673.
44. Livingstone. Personalised nutrition advice reduces intake of discretionary foods and beverages: findings from the Food4Me randomised controlled trial. The international journal of behavioral nutrition and physical activity. 2021. 10.1186/s12966-021-01136-5.
45. Taksler. Effect of Individualized Preventive Care Recommendations vs Usual Care on Patient Interest and Use of Recommendations: A Pilot Randomized Clinical Trial. JAMA network open. 2021. 10.1001/jamanetworkopen.2021.31455.
46. Fischer. Individualized Fluid Management Using the Pleth Variability Index: A Randomized Clinical Trial. Anesthesiology. 2020. 10.1097/ALN.0000000000003260..
47. Fernandez-Bustamante. Individualized PEEP to optimise respiratory mechanics during abdominal surgery: a pilot randomised controlled trial. British journal of anaesthesia. 2020. 10.1016/j.bja.2020.06.030.
48. Zheng. Personalized antiplatelet therapy guided by a novel detection of platelet aggregation function in stable coronary artery disease patients undergoing percutaneous coronary intervention: a randomized controlled clinical trial. European heart journal. Cardiovascular pharmacotherapy. 2020. 10.1093/ehjcvp/pvz059.
49. Doyle. Effect of personalised, mobile-accessible discharge instructions for patients leaving the emergency department: A randomised controlled trial. Emergency medicine Australasia : EMA. 2020. 10.1111/1742-6723.13516.
50. Booker. The effectiveness of an individualized sleep and shift work education and coaching program to manage shift work disorder in nurses: a randomized controlled trial. Journal of clinical sleep medicine : JCSM : official publication of the American Academy of Sleep Medicine. 2022. 10.5664/jcsm.9782.
51. Riese. Personalized ESM monitoring and feedback to support psychological treatment for depression: a pragmatic randomized controlled trial (Therap-i). BMC psychiatry. 2021. 10.1186/s12888-021-03123-3.
52. Vadiveloo. Effect of Personalized Incentives on Dietary Quality of Groceries Purchased: A Randomized Crossover Trial. JAMA network open. 2021. 10.1001/jamanetworkopen.2020.30921.
53. Sargent. An Individualized Intervention Increases Sleep Duration in Professional Athletes. Journal of strength and conditioning research. 2021. 10.1519/JSC.0000000000004138.
54. Jones. Effects of personalized depression prevention on anxiety through 18-month follow-up: A randomized controlled trial. Behaviour research and therapy. 2022. 10.1016/j.brat.2022.104156.

55. Luo. A simplified protocol for individualized regional citrate anticoagulation for hemodialysis: A single-center, randomized clinical study. Medicine. 2021. 10.1097/MD.0000000000024639.
56. Yen. Randomized Controlled Trial of Personalized Colorectal Cancer Risk Assessment vs Education to Promote Screening Uptake. The American journal of gastroenterology. 2021. 10.14309/ajg.0000000000000963.
57. Otsuki. Individualized nutritional treatment for acute stroke patients with malnutrition risk improves functional independence measurement: A randomized controlled trial. Geriatrics & gerontology international. 2020. 10.1111/ggi.13854.
58. Sauer-Zavala. A SMART approach to personalized care: preliminary data on how to select and sequence skills in transdiagnostic CBT. Cognitive behaviour therapy. 2022. 10.1080/16506073.2022.2053571.
59. Hartmann. Personalized collection of plasma from healthy donors: A randomized controlled trial of a novel technology-enabled nomogram. Transfusion. 2021. 10.1111/trf.16389.
60. Kirton. The Effects of Standardised versus Individualised Aerobic Exercise Prescription on Fitness-Fatness Index in Sedentary Adults: A Randomised Controlled Trial. Journal of sports science & medicine. 2022. 10.52082/jssm.2022.347.
61. Gao. Face-to-Face Instruction and Personalized Regimens Improve the Quality of Inpatient Bowel Preparation for Colonoscopy. Digestive diseases and sciences. 2022. 10.1007/s10620-021-07290-x..
62. Cheng. Outcomes of individualized goal-directed therapy based on cerebral oxygen balance in high-risk patients undergoing cardiac surgery: A randomized controlled trial. Journal of clinical anesthesia. 2020. 10.1016/j.jclinane.2020.110032.
63. Li. Preoperative carbohydrate loading with individualized supplemental insulin in diabetic patients undergoing gastrointestinal surgery: A randomized trial. International journal of surgery (London, England). 2022. 10.1016/j.ijsu.2021.106215.
64. Mandigout. Effect of individualized coaching at home on walking capacity in subacute stroke patients: A randomized controlled trial (Ticaa'dom). Annals of physical and rehabilitation medicine. 2021. 10.1016/j.rehab.2020.11.001.
65. Ishihara. Individualized follitropin delta dosing reduces OHSS risk in Japanese IVF/ICSI patients: a randomized controlled trial. Reproductive biomedicine online. 2021. 10.1016/j.rbmo.2021.01.023.
66. Arrieta. Effects of an individualized and progressive multicomponent exercise program on blood pressure, cardiorespiratory fitness, and body composition in long-term care residents: Randomized controlled trial. Geriatric nursing (New York, N.Y.). 2022. 10.1016/j.gerinurse.2022.03.005.
67. Schöttker. Protocol of the VICTORIA study: personalized vitamin D supplementation for reducing or preventing fatigue and enhancing quality of life of patients with colorectal tumor -Â randomized intervention trial. BMC cancer. 2020. 10.1186/s12885-020-07219-z.
68. Liu. Individualized PEEP ventilation between tumor resection and dural suture in craniotomy. Clinical neurology and neurosurgery. 2020. 10.1016/j.clineuro.2020.106027.
69. Vance. Can Individualized-Targeted Computerized Cognitive Training Benefit Adults with HIV-Associated Neurocognitive Disorder? The Training on Purpose Study (TOPS). AIDS and behavior. 2021. 10.1007/s10461-021-03230-y.
70. Taylor. Personalised yoga for burnout and traumatic stress in junior doctors. Postgraduate medical journal. 2020. 10.1136/postgradmedj-2019-137413.
71. Wang. Effect of an individualized digital coaching program on swallowing function in stroke patients. Acta neurologica Belgica. 2022. 10.1007/s13760-022-02153-2.
72. Morris. Randomized Trial of Individualized Texting for Adherence Building (iTAB) Plus Motivational Interviewing for PrEP Adherence in Transgender Individuals: The iM-PrEPT Study. Journal of acquired immune deficiency syndromes (1999). 2022. 10.1097/QAI.0000000000003091.
73. Vance. Can individualized-targeted computerized cognitive training improve everyday functioning in adults with HIV-associated neurocognitive disorder? Applied neuropsychology. Adult. 2022. 10.1080/23279095.2021.1906678.

74. Dutta. Efficacy of individualized homeopathic medicines in intervening with the progression of pre-hypertension to hypertension: A double-blind, randomized, placebo-controlled trial. Explore (New York, N.Y.). 2022. 10.1016/j.explore.2021.05.007.
75. Toğaç. Effects of preoperative individualized audiovisual education on anxiety and comfort in patients undergoing laparoscopic cholecystectomy: randomised controlled study. Patient education and counseling. 2021. 10.1016/j.pec.2020.08.026.
76. Griffin. Arthroscopic hip surgery compared with personalised hip therapy in people over 16 years old with femoroacetabular impingement syndrome: UK FASHIoN RCT. Health technology assessment (Winchester, England). 2022. 10.3310/FXII0508.
77. Misra. Individualized Homeopathic Medicines in Chronic Rhinosinusitis: Randomized, Double-Blind, Placebo-Controlled Trial. Homeopathy : the journal of the Faculty of Homeopathy. 2021. 10.1055/s-0040-1715842.
78. Santos Cerqueira. Effects of Individualized Ischemic Preconditioning on Protection Against Eccentric Exercise-Induced Muscle Damage: A Randomized Controlled Trial. Sports health. 2021. 10.1177/1941738121995414.
79. Hides. Telephone-based motivational interviewing enhanced with individualised personality-specific coping skills training for young people with alcohol-related injuries and illnesses accessing emergency or rest/recovery services: a randomized controlled trial (QuikFix). Addiction (Abingdon, England). 2021. 10.1111/add.15146.
80. Kontos. A Randomized Controlled Trial of Precision Vestibular Rehabilitation in Adolescents following Concussion: Preliminary Findings. The Journal of pediatrics. 2021. 10.1016/j.jpeds.2021.08.032.
81. Cureton. Randomised study within a trial (SWAT) to evaluate personalised versus standard text message prompts for increasing trial participant response to postal questionnaires (PROMPTS). Trials. 2021. 10.1186/s13063-021-05452-w.
82. Suen. Effect of brief, personalized feedback derived from momentary data on the mental health of women with risk of common mental disorders in Hong Kong: A randomized clinical trial. Psychiatry research. 2022. 10.1016/j.psychres.2022.114880.
83. Quan. Individualized Human Milk Fortification to Improve the Growth of Hospitalized Preterm Infants. Nutrition in clinical practice : official publication of the American Society for Parenteral and Enteral Nutrition. 2020. 10.1002/ncp.10366.
84. IsHak. Personalized treatments for depressive symptoms in patients with advanced heart failure: A pragmatic randomized controlled trial. PloS one. 2021. 10.1371/journal.pone.0244453.
85. Wonders. Cost-Savings Analysis of an Individualized Exercise Oncology Program in Early-Stage Breast Cancer Survivors: A Randomized Clinical Control Trial. JCO oncology practice. 2022. 10.1200/OP.21.00690.
86. Zenun Franco. Effectiveness of Web-Based Personalized Nutrition Advice for Adults Using the eNutri Web App: Evidence From the EatWellUK Randomized Controlled Trial. Journal of medical Internet research. 2022. 10.2196/29088.
87. Guo. Effects of an individualized analgesia protocol on the need for medical interventions after adenotonsillectomy in children: a randomized controlled trial. BMC anesthesiology. 2021. 10.1186/s12871-021-01263-3.
88. Cui. Precision implementation of early ambulation in elderly patients undergoing off-pump coronary artery bypass graft surgery: a randomized-controlled clinical trial. BMC geriatrics. 2020. 10.1186/s12877-020-01823-1.
89. Hogue. Personalized Blood Pressure Management During Cardiac Surgery With Cerebral Autoregulation Monitoring: A Randomized Trial. Seminars in thoracic and cardiovascular surgery. 2021. 10.1053/j.semtcvs.2020.09.032.

90. Htet. Rationale and design of a randomised controlled trial testing the effect of personalised diet in individuals with pre-diabetes or type 2 diabetes mellitus treated with metformin. BMJ open. 2020. 10.1136/bmjopen-2020-037859.
91. Yaremko. Functional Lung Avoidance for Individualized Radiation Therapy: Results of a Double-Masked, Randomized Controlled Trial. International journal of radiation oncology, biology, physics. 2022. 10.1016/j.ijrobp.2022.04.047.
92. Ingalls. Towards precision home visiting: results at six months postpartum from a randomized pilot implementation trial to assess the feasibility of a precision approach to Family Spirit. BMC pregnancy and childbirth. 2022. 10.1186/s12884-022-05057-4.
93. Siltanen. Effects of an Individualized Active Aging Counseling Intervention on Mobility and Physical Activity: Secondary Analyses of a Randomized Controlled Trial. Journal of aging and health. 2020. 10.1177/0898264320924258.
94. Zagorscak. How individuals change during internet-based interventions for depression: A randomized controlled trial comparing standardized and individualized feedback. Brain and behavior. 2020. 10.1002/brb3.1484.
95. Wang. Does providing personalized depression risk information lead to increased psychological distress and functional impairment? Results from a mixed-methods randomized controlled trial. Psychological medicine. 2022. 10.1017/S0033291720003955.
96. Perreault. Individualized high dairy proteinÂ +Â walking program supports bone health in pregnancy: a randomized controlled trial. The American journal of clinical nutrition. 2022. 10.1093/ajcn/nqac182.
97. Hoevenaars. Evaluation of Food-Intake Behavior in a Healthy Population: Personalized vs. One-Size-Fits-All. Nutrients. 2020. 10.3390/nu12092819.
98. Bock. Effect of Individualized Oral Health Care Training Provided to 6-16-Year-Old Psychiatric In-Patients-Randomized Controlled Study. International journal of environmental research and public health. 2022. 10.3390/ijerph192315615.
99. Nuijten. Evaluating the Impact of Adaptive Personalized Goal Setting on Engagement Levels of Government Staff With a Gamified mHealth Tool: Results From a 2-Month Randomized Controlled Trial. JMIR mHealth and uHealth. 2022. 10.2196/28801.
100. Robbins. Evaluating the impact of a sleep health education and a personalised smartphone application on sleep, productivity and healthcare utilisation among employees: results of a randomised clinical trial. BMJ open. 2022. 10.1136/bmjopen-2022-062121.
101. Zhang. Effects of personalized swallowing rehabilitation in patients with oral cancer after free flap transplantation: A cluster randomized controlled trial. Oral oncology. 2022. 10.1016/j.oraloncology.2022.106097.
102. Alemdar. The Effect of Individualized Developmental Care Practices in Preterm Infants. Complementary Medicine Research. 2020. 10.1159/000504357.
103. Hogan. HOME2 Study: Household Versus Personalized Decolonization in Households of Children With Methicillin-Resistant Staphylococcus aureus Skin and Soft Tissue Infection-A Randomized Clinical Trial. Clinical infectious diseases : an official publication of the Infectious Diseases Society of America. 2021. 10.1093/cid/ciaa752.
104. Wang. Individualized positive end-expiratory pressure with and without recruitment maneuvers in obese patients during bariatric surgery. The Kaohsiung journal of medical sciences. 2022. 10.1002/kjm2.12576.
105. Huibers. Hospital physicians' and older patients' agreement with individualised STOPP/START-based medication optimisation recommendations in a clinical trial setting. European geriatric medicine. 2022. 10.1007/s41999-022-00633-5.
106. Girrbach. Individualised positive end-expiratory pressure guided by electrical impedance tomography for robot-assisted laparoscopic radical prostatectomy: a prospective, randomised controlled clinical trial. British journal of anaesthesia. 2020. 10.1016/j.bja.2020.05.041.

107. Shimamoto. Providing Brief Personalized Therapies for Insomnia Among Workers Using a Sleep Prompt App: Randomized Controlled Trial. Journal of medical Internet research. 2022. 10.2196/36862.
108. Wynn. Personalized Reminders for Immunization Using Short Messaging Systems to Improve Human Papillomavirus Vaccination Series Completion: Parallel-Group Randomized Trial. JMIR mHealth and uHealth. 2021. 10.2196/26356.
109. Li. The Nursing Effect of Individualized Management on Patients With Diabetes Mellitus Type 2 and Hypertension. Frontiers in endocrinology. 2022. 10.3389/fendo.2022.846419.
110. Paton. Personalised reprogramming to prevent progressive pacemaker-related left ventricular dysfunction: A phase II randomised, controlled clinical trial. PloS one. 2021. 10.1371/journal.pone.0259450.
111. Applegate. Project ACTIVE: a Randomized Controlled Trial of Personalized and Patient-Centered Preventive Care in an Urban Safety-Net Setting. Journal of general internal medicine. 2021. 10.1007/s11606-020-06359-z.
112. Garey. Personalized Feedback for Smoking and Anxiety Sensitivity: A Randomized Controlled Trial. Substance use & misuse. 2021. 10.1080/10826084.2021.1900255.
113. Das. Efficacy of individualized homeopathic medicines in irritable bowel syndrome: A double-blind randomized, placebo-controlled trial. Explore (New York, N.Y.). 2022. 10.1016/j.explore.2022.10.004.
114. Zhao. Effects of prenatal individualized mixed management on breastfeeding and maternal health at three days postpartum: A randomized controlled trial. Early human development. 2020. 10.1016/j.earlhumdev.2019.104944.
115. Noguchi. A randomized phase III trial of personalized peptide vaccination for castrationâ€‘resistant prostate cancer progressing after docetaxel. Oncology reports. 2021. 10.3892/or.2020.7847.
116. Murphy. How Maternal BMI Modifies the Impact of Personalized Asthma Management in Pregnancy. J Allergy Clin Immunol Pract. 2020. 10.1016/j.jaip.2019.06.033.
117. He. Early individualized positive end-expiratory pressure guided by electrical impedance tomography in acute respiratory distress syndrome: a randomized controlled clinical trial. Critical care (London, England). 2021. 10.1186/s13054-021-03645-y.
118. Maïer. Effect of an individualized versus standard blood pressure management during mechanical thrombectomy for anterior ischemic stroke: the DETERMINE randomized controlled trial. Trials. 2022. 10.1186/s13063-022-06538-9.
119. Ma. A randomized clinical trial of genetic testing and personalized risk counselling in patients with type 2 diabetes receiving integrated care -The genetic testing and patient empowerment (GEM) trial. Diabetes research and clinical practice. 2022. 10.1016/j.diabres.2022.109969.
120. Yoo. Effect of structured individualized education on continuous glucose monitoring use in poorly controlled patients with type 1 diabetes: A randomized controlled trial. Diabetes research and clinical practice. 2022. 10.1016/j.diabres.2022.109209.
121. Hansen. Achilles tendon gait dynamics after rupture: A three-armed randomized controlled trial comparing an individualized treatment algorithm vs. operative or non-operative treatment. Foot and ankle surgery : official journal of the European Society of Foot and Ankle Surgeons. 2022. 10.1016/j.fas.2022.12.006.
122. Kuznia. Efficacy and Safety of a Personalized Vitamin D(3) Loading Dose Followed by Daily 2000 IU in Colorectal Cancer Patients with Vitamin D Insufficiency: Interim Analysis of a Randomized Controlled Trial. Nutrients. 2022. 10.3390/nu14214546.
123. Broers. Personalized eHealth Program for Life-style Change: Results From the "Do Cardiac Health Advanced New Generated Ecosystem (Do CHANGE 2)" Randomized Controlled Trial. Psychosomatic medicine. 2020. 10.1097/PSY.0000000000000802.
124. Paulus. Computer-delivered personalized feedback intervention for hazardous drinkers with elevated anxiety sensitivity: A pilot randomized controlled trial. Behaviour research and therapy. 2021. 10.1016/j.brat.2021.103847.

125. Li. Effect of individualized medical nutrition guidance on pregnancy outcomes in older pregnant women. The Journal of international medical research. 2021. 10.1177/03000605211033193.
126. Schweda. Implementation and evaluation of an individualized physical exercise promotion program in people with manifested risk factors for multimorbidity (MultiPill-Exercise): a study protocol for a pragmatic randomized controlled trial. BMC public health. 2022. 10.1186/s12889-022-13400-9.
127. Maas. Design of the PERSPECTIVE study: PERsonalized SPEeCh Therapy for actIVE conversation in Parkinson's disease (randomized controlled trial). Trials. 2022. 10.1186/s13063-022-06160-9.
128. Ilboudo. Effect of Personalized Support at Home on the Prevalence of Anemia in Pregnancy in Burkina Faso: A Cluster Randomized Trial. The American journal of tropical medicine and hygiene. 2021. 10.4269/ajtmh.20-1043.
129. Li. Clinical research on individualized hemodialysis preventing unconventional hypotension in diabetic nephropathy patient. The International journal of artificial organs. 2020. 10.1177/0391398819882697.
130. Hyatt. i-Move, a personalised exercise intervention for patients with advanced melanoma receiving immunotherapy: a randomised feasibility trial protocol. BMJ open. 2020. 10.1136/bmjopen-2019-036059.
131. Williams. What is the effectiveness of a personalised video story after an online diabetes risk assessment? A Randomised Controlled Trial. PloS one. 2022. 10.1371/journal.pone.0264749.
132. Ghosh. Individualized Homeopathic Medicines in Treatment of Hyperuricemia: Evaluation by Double-Blind, Randomized, Placebo-Controlled Trial. Homeopathy : the journal of the Faculty of Homeopathy. 2022. 10.1055/s-0042-1751272.
133. Keyu. A 3D printing personalized percutaneous puncture guide access plate for percutaneous nephrolithotomy: a pilot study. BMC urology. 2021. 10.1186/s12894-021-00945-x.
134. Biagioni. Adjunctive IgM-enriched immunoglobulin therapy with a personalised dose based on serum IgM-titres versus standard dose in the treatment of septic shock: a randomised controlled trial (IgM-fat trial) BMJ open. 2021. 10.1136/bmjopen-2019-036616.
135. Hsiao. The Preliminary Efficacy of a Sleep Self-management Intervention Using a Personalized Health Monitoring Device during Pregnancy. Behavioral sleep medicine. 2021. 10.1080/15402002.2020.1851230.
136. Bughin. Effects of an individualized exercise training program on severity markers of obstructive sleep apnea syndrome: a randomised controlled trial. Sleep medicine. 2020. 10.1016/j.sleep.2020.02.008.
137. Lorenzini. Personalized Telerehabilitation for a Head-mounted Low Vision Aid: A Randomized Feasibility Study. Optometry and vision science : official publication of the American Academy of Optometry. 2021. 10.1097/OPX.0000000000001704.
138. Dey. A Randomized, Double-Blind, Placebo-Controlled, Pilot Trial of Individualized Homeopathic Medicines for Cutaneous Warts. Homeopathy : the journal of the Faculty of Homeopathy. 2021. 10.1055/s-0040-1722232.
139. Smith. Four Conversations: A Randomized Controlled Trial of an Online, Personalized Coping and Decision Aid for Metastatic Breast Cancer Patients. Journal of palliative medicine. 2020. 10.1089/jpm.2019.0234.
140. Wang. The impact of providing personalized depression risk information on self-help and help-seeking behaviors: Results from a mixed methods randomized controlled trial. Depression and anxiety. 2021. 10.1002/da.23192.
141. Hansen. Individualized treatment for acute Achilles tendon rupture based on the Copenhagen Achilles Rupture Treatment Algorithm (CARTA): a study protocol for a multicenter randomized controlled trial. Trials. 2020. 10.1186/s13063-020-04332-z.
142. Wang. Personalized nutrition intervention improves nutritional status and quality of life of colorectal cancer survivors in the community: A randomized controlled trial. Nutrition (Burbank, Los Angeles County, Calif.). 2022. 10.1016/j.nut.2022.111835.

143. Teeters. A randomized pilot trial of a mobile phone-based brief intervention with personalized feedback and interactive text messaging to reduce driving after cannabis use and riding with a cannabis impaired driver. Journal of substance abuse treatment. 2022. 10.1016/j.jsat.2022.108867.
144. Wang. Intraoperative right heart function with individualized mechanical ventilation in laparoscopic surgery with Trendelenburg positioning: A randomized-controlled study. Heart & lung : the journal of critical care. 2022. 10.1016/j.hrtlng.2022.12.007.
145. Lo. Effects of Individualized Aerobic Exercise Training on Physical Activity and Health-Related Physical Fitness among Middle-Aged and Older Adults with Multimorbidity: A Randomized Controlled Trial. International journal of environmental research and public health. 2020. 10.3390/ijerph18010101.
146. Kukralova. The Impact of Individualized Hemodynamic Management on Intraoperative Fluid Balance and Hemodynamic Interventions during Spine Surgery in the Prone Position: A Prospective Randomized Trial. Medicina (Kaunas, Lithuania). 2022. 10.3390/medicina58111683.
147. Broers. A Personalized eHealth Intervention for Lifestyle Changes in Patients With Cardiovascular Disease: Randomized Controlled Trial. Journal of medical Internet research. 2020. 10.2196/14570.
148. Schneider. BRE12-158: A Postneoadjuvant, Randomized Phase II Trial of Personalized Therapy Versus Treatment of Physician's Choice for Patients With Residual Triple-Negative Breast Cancer. Journal of clinical oncology : official journal of the American Society of Clinical Oncology. 2022. 10.1200/JCO.21.01657.
149. Błajda. Application of Personalized Education in the Mobile Medical App for Breast Self-Examination. International journal of environmental research and public health. 2022. 10.3390/ijerph19084482.
150. Shimada. Effect of individualized occupational therapy on social functioning in patients with schizophrenia: A five-year follow-up of a randomized controlled trial. Journal of psychiatric research. 2022. 10.1016/j.jpsychires.2022.10.066.
151. Magits. Comparing the Outcomes of a Personalized Versus Nonpersonalized Home-Based Auditory Training Program for Cochlear Implant Users. Ear and hearing. 2022. 10.1097/AUD.0000000000001295.
152. Reinders. Effectiveness and cost-effectiveness of personalised dietary advice aiming at increasing protein intake on physical functioning in community-dwelling older adults with lower habitual protein intake: rationale and design of the PROMISS randomised controlled trial. BMJ open. 2020. 10.1136/bmjopen-2020-040637.
153. Tandon. Heal-me PiONEer (personalized online nutrition and exercise): An RCT assessing 2 levels of app-based programming in individuals with chronic disease. Contemporary clinical trials. 2022. 10.1016/j.cct.2022.106791.
154. Buckner. On-line personalized feedback intervention for negative affect and cannabis: A pilot randomized controlled trial. Experimental and clinical psychopharmacology. 2020. 10.1037/pha0000304.
155. Purper-Ouakil. Personalized at-home neurofeedback compared to long-acting methylphenidate in children with ADHD: NEWROFEED, a European randomized noninferiority trial. Journal of child psychology and psychiatry, and allied disciplines. 2022. 10.1111/jcpp.13462.
156. Hanssen. An ecological momentary intervention incorporating personalised feedback to improve symptoms and social functioning in schizophrenia spectrum disorders. Psychiatry research. 2020. 10.1016/j.psychres.2019.112695.
157. Hegelund. The impact of a personalised action plan delivered at discharge to patients with COPD on readmissions: a pilot study. Scandinavian journal of caring sciences. 2020. 10.1111/scs.12798.
158. Heinemann. Benefit of Digital Tools Used for Integrated Personalized Diabetes Management: Results From the PDM-ProValue Study Program. Journal of diabetes science and technology. 2020. 10.1177/1932296819867686.
159. Trent. Using Innovation to Address Adolescent and Young Adult Health Disparities in Pelvic Inflammatory Disease: Design of the Technology Enhanced Community Health Precision Nursing (TECH-PN) Trial. The Journal of infectious diseases. 2021. 10.1093/infdis/jiab157.

160. Chang. Individualized intervention for frail non-dialysis elderly patients with chronic kidney disease: protocol for a randomized controlled trial. BMC geriatrics. 2020. 10.1186/s12877-020-1491-6.
161. Huang. Efficacy of individualized education in patients with type 2 diabetes mellitus: A randomized clinical study protocol. Medicine. 2020. 10.1097/MD.0000000000023625.
162. Zhao. An evaluation of a prenatal individualised mixed management intervention addressing breastfeeding outcomes and postpartum depression: A ramdomised controlled trial. Journal of clinical nursing. 2021. 10.1111/jocn.15684.
163. Chapman. Personalised Adherence Support for Maintenance Treatment of Inflammatory Bowel Disease: A Tailored Digital Intervention to Change Adherence-related Beliefs and Barriers. Journal of Crohn's & colitis. 2020. 10.1093/ecco-jcc/jjz034.
164. Jindal. Randomized controlled trial of individualized, low dose, fixed duration lenalidomide maintenance versus observation after frontline chemo-immunotherapy in CLL. Leukemia & Lymphoma. 2021. 10.1080/10428194.2021.1885668.
165. Neumeier. POWERSforID: Personalized online weight and exercise response system for individuals with intellectual disability: A randomized controlled trial. Disability and health journal. 2021. 10.1016/j.dhjo.2021.101111.
166. Woisard. How a personalised transportable folding device for seating impacts dysphagia. European archives of oto-rhino-laryngology : official journal of the European Federation of Oto-Rhino-Laryngological Societies (EUFOS) : affiliated with the German Society for Oto-Rhino-Laryngology - Head and Neck Surgery. 2020. 10.1007/s00405-019-05657-5.
167. Schafer. A block randomised controlled trial investigating changes in postural control following a personalised 12-week exercise programme for individuals with lower limb amputation. Gait & posture. 2021. 10.1016/j.gaitpost.2020.12.001.
168. Konerding. A pragmatic randomised controlled trial referring to a Personalised Self-management SUPport Programme (P-SUP) for persons enrolled in a disease management programme for type 2 diabetes mellitus and/or for coronary heart disease. Trials. 2021. 10.1186/s13063-021-05636-4.
169. Zhang. Angiotensin II Regulates the Neural Expression of Subjective Fear in Humans: A Precision Pharmaco-Neuroimaging Approach. Biological psychiatry. Cognitive neuroscience and neuroimaging. 2022. 10.1016/j.bpsc.2022.09.008.
170. Tariq. A Last-Ditch Effort and Personalized Surgeon Letter Improves PROMs Follow-Up Rate in Sports Medicine Patients: A Crossover Randomized Controlled Trial. J Knee Surgery. 2020. 10.1055/s-0039-1694057.
171. Parewa. Individualized Homeopathic Medicines in the Treatment of Generalized Anxiety Disorder: A Double-Blind, Randomized, Placebo-Controlled, Pilot Trial. Complementary Medicine Research. 2021. 10.1159/000514524.
172. Cioe. Personalized feedback improves cardiovascular risk perception and physical activity levels in persons with HIV: results of a pilot randomized clinical trial. AIDS Care. 2020. 10.1080/09540121.2021.1874271.
173. Angosta. Incorporating Social Networks and Event-Specific Information in a Personalized Feedback Intervention to Reduce Drinking Among Young Adults. Alcohol and alcoholism (Oxford, Oxfordshire). 2022. 10.1093/alcalc/agac005.
174. Wu. An alternative method for personalized tourniquet pressure in total knee arthroplasty: a prospective randomized and controlled study. Scientific reports. 2022. 10.1038/s41598-022-13672-6.
175. Boyle. A Gamified Personalized Normative Feedback App to Reduce Drinking Among Sexual Minority Women: Randomized Controlled Trial and Feasibility Study. Journal of medical Internet research. 2022. 10.2196/34853.
176. Siemens. Individualized, Intraoperative Dosing of Fibrinogen Concentrate for the Prevention of Bleeding in Neonatal and Infant Cardiac Surgery Using Cardiopulmonary Bypass (FIBCON): A Phase

1b/2a Randomized Controlled Trial. Circulation. Cardiovascular interventions. 2020. 10.1161/CIRCINTERVENTIONS.120.009465.
177. Giovannetti. Impact of an individualized and adaptive cognitive intervention on working memory, planning and fluid reasoning processing in preschoolers from poor homes. Child neuropsychology : a journal on normal and abnormal development in childhood and adolescence. 2022. 10.1080/09297049.2021.1998406.
178. Wachter. Angiotensin receptor neprilysin inhibition versus individualized RAAS blockade: design and rationale of the PARALLAX trial. ESC heart failure. 2020. 10.1002/ehf2.12694.
179. Kastelz. Personalized physical rehabilitation program and employment in kidney transplant recipients: a randomized trial. Transplant international : official journal of the European Society for Organ Transplantation. 2021. 10.1111/tri.13868.
180. Politi. A Randomized Controlled Trial Evaluating the BREASTChoice Tool for Personalized Decision Support About Breast Reconstruction After Mastectomy. Annals of surgery. 2020. 10.1097/SLA.0000000000003444.
181. Litt. Individualized assessment and treatment program (IATP) for cannabis use disorder: Randomized controlled trial with and without contingency management. Psychology of addictive behaviors : journal of the Society of Psychologists in Addictive Behaviors. 2020. 10.1037/adb0000491.
182. Stapleton. A randomized controlled trial of a web-based personalized feedback intervention targeting frequent indoor tanning bed users: Engagement, acceptability, and preliminary behavioral outcomes. Journal of health psychology. 2022. 10.1177/1359105320982038.
183. Rollo. Impact on Dietary Intake of Two Levels of Technology-Assisted Personalized Nutrition: A Randomized Trial. Nutrients. 2020. 10.3390/nu12113334.
184. Arntzen. Group-based, individualized, comprehensive core stability and balance intervention provides immediate and long-term improvements in walking in individuals with multiple sclerosis: A randomized controlled trial. Physiotherapy research international : the journal for researchers and clinicians in physical therapy. 2020. 10.1002/pri.1798.
185. Frith. Pragmatic randomised controlled trial of a personalised intervention for carers of people requiring home oxygen therapy. Chronic respiratory disease. 2020. 10.1177/1479973119897277.
186. Zhang. Individualized positive end-expiratory pressure in patients undergoing thoracoscopic lobectomy: a randomized controlled trial. Brazilian journal of anesthesiology (Elsevier). 2021. 10.1016/j.bjane.2021.04.001.
187. Moreno-Peral. Use of a personalised depression intervention in primary care to prevent anxiety: a secondary study of a cluster randomised trial. The British journal of general practice : the journal of the Royal College of General Practitioners. 2021. 10.3399/bjgp20X714041.
188. Hummer. Cognitive-Affective Change Mechanisms in Personalized Normative Feedback via the Articulated Thoughts in Simulated Situations Paradigm. International journal of environmental research and public health. 2020. 10.3390/ijerph17030690.
189. Almabadi. The Effect of a Personalized Oral Health Education Program on Periodontal Health in an At-Risk Population: A Randomized Controlled Trial. International journal of environmental research and public health. 2021. 10.3390/ijerph18020846.
190. Zupkauskiene. Changes in health-related quality of life, motivation for physical activity, and the levels of anxiety and depression after individualized aerobic training in subjects with metabolic syndrome. Hellenic journal of cardiology : HJC = Hellenike kardiologike epitheorese. 2022. 10.1016/j.hjc.2022.04.003.
191. Oh. Effect of personalized extracorporeal biofeedback device for pelvic floor muscle training on urinary incontinence after robot-assisted radical prostatectomy: A randomized controlled trial. Neurourology and urodynamics. 2020. 10.1002/nau.24247.

192. Clemons. A randomized trial of individualized versus standard of care antiemetic therapy for breast cancer patients at high risk for chemotherapy-induced nausea and vomiting. Breast (Edinburgh, Scotland). 2020. 10.1016/j.breast.2020.11.002.
193. Díaz-Cambronero. Effect of an individualized versus standard pneumoperitoneum pressure strategy on postoperative recovery: a randomized clinical trial in laparoscopic colorectal surgery. The British journal of surgery. 2020. 10.1002/bjs.11736.
194. Gottlieb. Effects of In-Person Assistance vs Personalized Written Resources About Social Services on Household Social Risks and Child and Caregiver Health: A Randomized Clinical Trial. JAMA network open. 2020. 10.1001/jamanetworkopen.2020.0701.
195. Nayak. Efficacy of individualized homeopathy as an adjunct to standard of care of COVID-19: A randomized, single-blind, placebo-controlled study. Complementary therapies in clinical practice. 2022. 10.1016/j.ctcp.2022.101602.
196. Marshall. Impact of an online, individualised, patient reported outcome measures based patient decision aid on patient expectations, decisional regret, satisfaction, and health-related quality-of-life for patients considering total knee arthroplasty: Results from a randomised controlled trial. Journal of evaluation in clinical practice. 2022. 10.1111/jep.13804.
197. Martínez-Velilla. Recovery of the Decline in Activities of Daily Living After Hospitalization Through an Individualized Exercise Program: Secondary Analysis of a Randomized Clinical Trial. The journals of gerontology. Series A, Biological sciences and medical sciences. 2021. 10.1093/gerona/glab032.
198. Nuttall. A Blinded Randomized Trial Comparing Standard Activated Clotting Time Heparin Management to High Target Active Clotting Time and Individualized Hepcon HMS Heparin Management in Cardiopulmonary Bypass Cardiac Surgical Patients. Annals of thoracic and cardiovascular surgery : official journal of the Association of Thoracic and Cardiovascular Surgeons of Asia. 2022. 10.5761/atcs.oa.21-00222.
199. Panwar. Standard care versus individualized blood pressure targets among critically ill patients with shock: A multicenter feasibility and preliminary efficacy study. Journal of critical care. 2022. 10.1016/j.jcrc.2022.154052.
200. Pellicori. Effects of spironolactone on serum markers of fibrosis in people at high risk of developing heart failure: rationale, design and baseline characteristics of a proof-of-concept, randomised, precision-medicine, prevention trial. The Heart OMics in AGing (HOMAGE) trial. European journal of heart failure. 2020. 10.1002/ejhf.1716.
201. Zimmer-Gembeck. The Circle of Security Parenting Program (COS-P): A Randomized Controlled Trial of a Low Intensity, Individualized Attachment-Based Program With at-Risk Caregivers. Behavior therapy. 2022. 10.1016/j.beth.2021.07.003.
202. Oh. Individualized education focusing on self-management improved the knowledge and self-management behaviour of elderly people with atrial fibrillation: A randomized controlled trial. International journal of nursing practice. 2021. 10.1111/ijn.12902.
203. Gan. Application of Personalized Navigation Templates to Oxford Single Condylar Replacement in a Chinese Population. J Knee Surgery. 2020. 10.1055/s-0040-1702188.
204. Barlesi. Comprehensive Genome Profiling in Patients With Metastatic Non-Small Cell Lung Cancer: The Precision Medicine Phase II Randomized SAFIR02-Lung/IFCT 1301 Trial. Clinical cancer research : an official journal of the American Association for Cancer Research. 2022. 10.1158/1078-0432.CCR-22-0371.
205. Manasse. The project REBOOT protocol: Evaluating a personalized inhibitory control training as an adjunct to cognitive behavioral therapy for bulimia nervosa and binge-eating disorder. The International journal of eating disorders. 2020. 10.1002/eat.23225.
206. Gierula. Personalized Rate-Response Programming Improves Exercise Tolerance After 6 Months in People With Cardiac Implantable Electronic Devices and Heart Failure: A Phase II Study. Circulation. 2020. 10.1161/CIRCULATIONAHA.119.045066.

207. Marin-Alejandre. Effects of two personalized dietary strategies during a 2-year intervention in subjects with nonalcoholic fatty liver disease: A randomized trial. Liver international : official journal of the International Association for the Study of the Liver. 2021. 10.1111/liv.14818.
208. Li. Effect of pressure-controlled ventilation-volume guaranteed mode combined with individualized positive end-expiratory pressure on respiratory mechanics, oxygenation and lung injury in patients undergoing laparoscopic surgery in Trendelenburg position. Journal of clinical monitoring and computing. 2022. 10.1007/s10877-021-00750-9.
209. Chatwin. An intelligent insole system with personalised digital feedback reduces foot pressures during daily life: An 18-month randomised controlled trial. Diabetes research and clinical practice. 2021. 10.1016/j.diabres.2021.109091.
210. Reinders. The cost effectiveness of personalized dietary advice to increase protein intake in older adults with lower habitual protein intake: a randomized controlled trial. European journal of nutrition. 2022. 10.1007/s00394-021-02675-0.
211. Wang. The efficacy of 3D personalized insoles in moderate adolescent idiopathic scoliosis: a randomized controlled trial. BMC musculoskeletal disorders. 2022. 10.1186/s12891-022-05952-z.
212. Heapy. Incorporating walking into cognitive behavioral therapy for chronic pain: safety and effectiveness of a personalized walking intervention. Journal of behavioral medicine. 2021. 10.1007/s10865-020-00193-8.
213. Rijnaarts. Increasing dietary fibre intake in healthy adults using personalised dietary advice compared with general advice: a single-blind randomised controlled trial. Public health nutrition. 2021. 10.1017/S1368980020002980.
214. Xu. Feasibility of Investigational Procedures and Efficacy of a Personalized Omega-3 Dietary Intervention in Alleviating Pain and Psychoneurological Symptoms in Breast Cancer Survivors. Pain management nursing : official journal of the American Society of Pain Management Nurses. 2022. 10.1016/j.pmn.2022.03.007.
215. Paudel. Impact of hospital pharmacist-delivered individualised pharmaceutical service intervention on clinical and patient-reported outcomes in patients with hypertension: a randomised controlled trial. European Journal of Hospital Pharmacy. 2020. 10.1136/ejhpharm-2020-002512.
216. Tahkola. Personalized text message and checklist support for initiation of antihypertensive medication: the cluster randomized, controlled check and support trial. Scandinavian journal of primary health care. 2020. 10.1080/02813432.2020.1753380.
217. Testa. Preventing College Sexual Victimization by Reducing Hookups: a Randomized Controlled Trial of a Personalized Normative Feedback Intervention. Prevention science : the official journal of the Society for Prevention Research. 2020. 10.1007/s11121-020-01098-3.
218. Luther. Mobile enhancement of motivation in schizophrenia: A pilot randomized controlled trial of a personalized text message intervention for motivation deficits. Journal of consulting and clinical psychology. 2020. 10.1037/ccp0000599.
219. Wu. Increasing Skin Cancer Prevention in Young Adults: the Cumulative Impact of Personalized UV Photography and MC1R Genetic Testing. Journal of cancer education : the official journal of the American Association for Cancer Education. 2022. 10.1007/s13187-022-02232-1.
220. Kwak. Findings From a Prospective Randomized Controlled Trial of an Individualized Music Listening Program for Persons With Dementia. Journal of applied gerontology : the official journal of the Southern Gerontological Society. 2020. 10.1177/0733464818778991.
221. Montone. Precision medicine versus standard of care for patients with myocardial infarction with non-obstructive coronary arteries (MINOCA): rationale and design of the multicentre, randomised PROMISE trial. EuroIntervention : journal of EuroPCR in collaboration with the Working Group on Interventional Cardiology of the European Society of Cardiology. 2022. 10.4244/EIJ-D-22-00178.

222. Lang. A randomized controlled trial investigating effects of an individualized pedometer driven walking program on chronic low back pain. BMC musculoskeletal disorders. 2021. 10.1186/s12891-021-04060-8.
223. He. The Effect of Health Education Combined with Personalized Psychological Nursing Intervention on Pregnancy Outcome of Pregnant Women with Gestational Diabetes Mellitus. BioMed research international. 2022. 10.1155/2022/3157986.
224. Nayak. Individualized homeopathic medicines and Urtica urens mother tincture in treatment of hyperuricemia: an open, randomized, pragmatic, pilot trial. J Complement Integr Med. 2020. 10.1515/jcim-2020-0129.
225. Gómez-Soria. Effectiveness of Personalized Cognitive Stimulation in Older Adults with Mild Possible Cognitive Impairment: A 12-month Follow-up Cognitive Stimulation in Mild Cognitive Impairment. Clinical gerontologist. 2022. 10.1080/07317115.2021.1937764.
226. Xu Q. Effects of dynamic individualized PEEP guided by driving pressure in laparoscopic surgery on postoperative atelectasis in elderly patients: a prospective randomized controlled trial. BMC anesthesiology. 2022. 10.1186/s12871-022-01613-9.
227. Wen. A Pilot Study on Approach Bias Modification in Smoking Cessation: Activating Personalized Alternative Activities for Smoking in the Context of Increased Craving. International journal of behavioral medicine. 2022. 10.1007/s12529-021-10033-x.
228. Rochow. Individualized target fortification of breast milk with protein, carbohydrates, and fat for preterm infants: A double-blind randomized controlled trial. Clinical nutrition (Edinburgh, Scotland). 2021. 10.1016/j.clnu.2020.04.031.
229. Di. Lack of effects of evidence-based, individualised counselling on medication use in insured patients with mild hypertension in China: a randomised controlled trial. BMJ evidence-based medicine. 2020. 10.1136/bmjebm-2019-111197.
230. Gautier. Impact of personalized text messages from pharmacists on medication adherence in type 2 diabetes in France: A real-world, randomized, comparative study. Patient education and counseling. 2021. 10.1016/j.pec.2021.02.022.
231. Zhang. Effects of individualized administration of folic acid on prothrombotic state and vascular endothelial function with H-type hypertension: A double-blinded, randomized clinical cohort study. Medicine. 2022. 10.1097/MD.0000000000028628.
232. Rodríguez-Torres. Effects of an Individualized Comprehensive Rehabilitation Program on Impaired Postural Control in Women With Chronic Pelvic Pain: A Randomized Controlled Trial. Arch Phys Med Rehabil. 2020. 10.1016/j.apmr.2020.02.019.
233. Syed. Implementing a sleep technician-supervised and personalized APAP interface fitting session prior to initiation of home APAP therapy improves adherence in patients with obstructive sleep apnea. Journal of clinical sleep medicine : JCSM : official publication of the American Academy of Sleep Medicine. 2021. 10.5664/jcsm.9368.
234. Zeng. A study for precision diagnosing and treatment strategies in difficult-to-treat AIDS cases and HIV-infected patients with highly fatal or highly disabling opportunistic infections: Study protocol for antiretroviral therapy timing in AIDS patients with toxoplasma encephalitis. Medicine. 2020. 10.1097/MD.0000000000021141.
235. Calvo. Can an individualized adherence education program delivered by nurses improve therapeutic adherence in elderly people with acute myocardial infarction?: A randomized controlled study. International journal of nursing studies. 2021. 10.1016/j.ijnurstu.2021.103975.
236. Weber. Effect of individualized PEEP titration guided by intratidal compliance profile analysis on regional ventilation assessed by electrical impedance tomography - a randomized controlled trial. BMC anesthesiology. 2020. 10.1186/s12871-020-00960-9.

237. Paterno. Feasibility of a pilot, randomized controlled trial using a personalized health monitoring device with pregnant women for behavioral sleep research. Applied nursing research : ANR. 2020. 10.1016/j.apnr.2020.151246.
238. Passaro. Personalized Cognitive Counseling Reduces Drinking Expectancy Among Men Who Have Sex with Men and Transgender Women in Lima, Peru: A Pilot Randomized Controlled Trial. AIDS and behavior. 2020. 10.1007/s10461-020-02882-6.
239. Held. 3D-printed individualized tooth-borne tissue retraction devices compared to conventional dental splints for head and neck cancer radiotherapy: a randomized controlled trial. Radiation oncology (London, England). 2021. 10.1186/s13014-021-01803-8.
240. Zhang. Task-related functional magnetic resonance imaging-based neuronavigation for the treatment of depression by individualized repetitive transcranial magnetic stimulation of the visual cortex. Science China. Life sciences. 2021. 10.1007/s11427-020-1730-5.
241. Kim. Effects of an Individualized Educational Program for Korean Patients With Chronic Low Back Pain: A Randomized Controlled Trial. The journal of nursing research : JNR. 2021. 10.1097/jnr.0000000000000455.
242. Mozafarinia. Effectiveness of a personalized health profile on specificity of self-management goals among people living with HIV in Canada: findings from a blinded pragmatic randomized controlled trial. Quality of life research : an international journal of quality of life aspects of treatment, care and rehabilitation. 2022. 10.1007/s11136-022-03245-5.
243. Piernas. Evaluation of an intervention to provide brief support and personalized feedback on food shopping to reduce saturated fat intake (PC-SHOP): A randomized controlled trial. PLoS medicine. 2020. 10.1371/journal.pmed.1003385.
244. Karabulut. A new method in robotic-assisted laparoscopic radical prostatectomy: personalised neuroprotective surgery with neuromonitoring system-randomised controlled study. International urology and nephrology. 2020. 10.1007/s11255-019-02295-y.
245. Marcu. Non-Steroidal Anti-Inflammatory Drug Etoricoxib Facilitates the Application of Individualized Exercise Programs in Patients with Ankylosing Spondylitis. Medicina (Kaunas, Lithuania). 2020. 10.3390/medicina56060270.
246. Patel. Tobacco cessation effects on oral health by group and individualized motivational therapy in 12 to 18 years old boys - A randomized controlled study. Journal of the Indian Society of Pedodontics and Preventive Dentistry. 2020. 10.4103/JISPPD.JISPPD_333_20.
247. Coulibaly. Personalized treatment supported by automated quantitative fluid analysis in active neovascular age-related macular degeneration (nAMD)-a phase III, prospective, multicentre, randomized study: design and methods. Eye (London, England). 2022. 10.1038/s41433-022-02154-8.
248. de Campos. An individualised self-management exercise and education program did not prevent recurrence of low back pain but may reduce care seeking: a randomised trial. Journal of physiotherapy. 2020. 10.1016/j.jphys.2020.06.006.
249. Xin. Cost-effectiveness of the PDSAFE personalised physiotherapy intervention for fall prevention in Parkinson's: an economic evaluation alongside a randomised controlled trial. BMC neurology. 2020. 10.1186/s12883-020-01852-8.
250. Chen. An Individualized, Interactive, and Advance Care Planning Intervention Promotes Transitions in Prognostic Awareness States Among Terminally Ill Cancer Patients in Their Last Six Months-A Secondary Analysis of a Randomized Controlled Trial. Journal of pain and symptom management. 2020. 10.1016/j.jpainsymman.2020.01.012.
251. Matthew. Effects of personalized music intervention on nurse burnout: A feasibility randomized controlled trial. Nursing & Health Sciences. 2022. 10.1111/nhs.12984.
252. Steffens. Individualised, targeted step count intervention following gastrointestinal cancer surgery: The Fit-4-Home randomised clinical trial. ANZ Journal of Surgery. 2021. 10.1111/ans.17212.

253. Vander Vegt. A Comparison of Generalized and Individualized Vestibular Rehabilitation Therapy in a Military TBI Sample. Journal of Head Trauma Rehabilitation. 2022. 10.1097/HTR.0000000000000777.
254. Rathe. Inflammatory effects of individualized abutments bonded onto titanium base on peri-implant tissue health: A randomized controlled clinical trial. Clinical Implant Dentistry and Related Research. 2021. 10.1111/cid.13050.
255. Kachmar. Personalized balance games for children with cerebral palsy: A pilot study. Journal of Pediatric Rehabilitation Medicine. 2021. 10.3233/PRM-190666.
256. Pytel. Personalized Repetitive Transcranial Magnetic Stimulation for Primary Progressive Aphasia. Journal of Alzheimer's Disease . 2021. 10.3233/JAD-210566.
257. Heij. Implementing a Personalized Physical Therapy Approach (Coach2Move) Is Effective in Increasing Physical Activity and Improving Functional Mobility in Older Adults: A Cluster-Randomized, Stepped Wedge Trial. Physical Therapy. 2022. 10.1093/ptj/pzac138.
258. Perna. The Potential of Personalized Virtual Reality in Palliative Care: A Feasibility Trial. American Journal of Hospice and Palliative Medicine. 2021. 10.1177/1049909121994299.
259. Laskar. Individualized Homeopathic Medicines in the Treatment of Tinea Corporis: Double-Blind, Randomized, Placebo-Controlled Trial. Homeopathy. 2022. 10.1055/s-0042-1750799.
260. Dutta. Efficacy of individualized homeopathic medicines in treatment of post-stroke hemiparesis: A randomized trial. EXPLORE. 2022. 10.1016/j.explore.2022.08.017.
261. Szilagyi. Effect of Personalized Messages Sent by a Health System's Patient Portal on Influenza Vaccination Rates: a Randomized Clinical Trial. Journal of General Internal Medicine. 2021. 10.1007/s11606-021-07023-w.
262. O'Dwyer. Personalized Biofeedback on Inhaler Adherence and Technique by Community Pharmacists: AÂ Cluster Randomized Clinical Trial. The Journal of Allergy and Clinical Immunology: In Practice. 2019. 10.1016/j.jaip.2019.09.008.
